# Inexpensive workflow for simultaneous monitoring of HIV viral load and detection of SARS-CoV-2 infection

**DOI:** 10.1101/2021.08.18.21256786

**Authors:** Gaurav K Gulati, Nuttada Panpradist, Samuel W A Stewart, Ingrid A Beck, Ceejay Boyce, Amy K Oreskovic, Claudia García-Morales, Santiago Avila-Ríos, Peter D. Han, Gustavo Reyes-Terán, Lea M. Starita, Lisa M Frenkel, Barry R Lutz, James J Lai

## Abstract

**Background:** COVID-19 pandemic interrupted routine care for individuals living with HIV, putting them at risk of becoming virologically unsuppressed and ill. Often they are at high risk for exposure to SARS-CoV-2 infection and severe disease once infected. For this population, it is urgent to closely monitor HIV plasma viral load (**VL**) and screen for SARS-COV-2 infection.

**Method:** We have developed a non-proprietary method to isolate RNA from plasma, nasal secretions (**NS**), or both. HIV, SARS-CoV-2, and human RP targets in extracted RNA are then RT-qPCR to estimate the VL and classify HIV/SARS-CoV-2 status (*i*.*e*., HIV as VL failure or suppressed; SARS-CoV-2 as positive, presumptive positive, negative, or indeterminate). We evaluated this workflow on 133 clinical specimens: 40 plasma specimens (30 HIV-seropositive), 67 NS specimens (31 SARS-CoV-2-positive), and 26 pooled plasma/NS specimens (26 HIV-positive with 10 SARS-CoV-2-positive), and compared the results obtained using the in-house extraction to those using a commercial extraction kit.

**Results:** In-house extraction had a detection limit of 200-copies/mL for HIV and 100-copies/mL for SARS-CoV-2. In-house and commercial methods yielded positively correlated HIV VL (R^2^: 0.98 for contrived samples; 0.81 for seropositive plasma). SARS-CoV-2 detection had 100% concordant classifications in contrived samples, and in clinical NS extracted by in-house method, excluding indeterminate results, was 95% concordant (25 positives, 6 presumptive positives, and 31 negatives) to those using the commercial method. Analysis of pooled plasma/NS showed R^2^ of 0.91 (contrived samples) and 0.71 (clinical specimens) for HIV VL correlations obtained by both extraction methods, while SARS-CoV-2 detection showed 100% concordance in contrived and clinical specimens.

**Interpretation:** Our low-cost workflow for molecular testing of HIV and SARS-CoV-2 could serve as an alternative to current standard assays for laboratories in low-resource settings.

## Introduction

COVID-19 pandemic has stalled public health responses to address other pre-existing global epidemics such as HIV^[1]^. People living with HIV (**PLH**) are impacted by the SARS-CoV-2/HIV syndemic in multiple ways^[2]^. Reportedly, PLH have fears for COVID-19 exposure during routine clinical care^[3, 4]^, which can impact HIV treatment and result in unsuppressed HIV replication. PLH have increased morbidity and mortality if infected with SARS-CoV-2^[5-10]^; especially those with detectable viremia and/or low CD4 counts^[11]^. A recent study reports that symptoms of acute HIV infection can be similar to SARS-CoV-2 symptoms^[12]^; thus testing may not be performed for HIV infection. Prolonged SARS-CoV-2 infections occur in immunocompromised individuals, including PLH, in whom multiple variants can evolve^[13]^. Consequently, early diagnosis of SARS-CoV-2 in PLH, to allow treatment and isolation, could benefit both the individual and public health. “Lockdowns” due to COVID-19 have limited transportation and reduced access to in-person doctor visits^[14]^, and have impacted the global economy, thus interfering the delivery of effective HIV diagnostics and treatments^[15]^.

“A call to action,”^[16]^ encouraging HIV researchers to innovate approaches to diminish the impact of COVID-19 on HIV treatment, led us to streamline a workflow to screen for SARS-CoV-2 infection while monitoring HIV RNA viral load (**HIV VL**). Standard laboratory-based molecular methods previously used to detect HIV RNA have been applied to detect SARS-CoV-2 RNA. These molecular tests traditionally involve “extraction” to concentrate and purify the RNA from inhibitors present in biological samples, followed by reverse-transcription quantitative polymerase chain reaction (**RT-qPCR**) to quantify viral loads. Patients hospitalized with COVID-19 reportedly tested positive for SARS-CoV-2 RNA in plasma^[17]^ and those with higher SARS-CoV-2 viremia were associated with worse clinical outcomes^[18, 19]^. Therefore, quantification of SARS-CoV-2 RNA in plasma may be a useful indicator for disease severity. Depending on the sensitivity of the RT-qPCR, plasma SARS-CoV-2 RNA was detected in 19-74% of patients diagnosed with SARS-CoV-2 via respiratory samples^[18-20]^. SARS-CoV-2 viral loads in respiratory samples can remain above 10^4^ copies/mL across six weeks after initial infection, whereas SARS-CoV-2 viral loads in the blood were approximately 500 copies/mL in the first two weeks and completely cleared after the third week of initial infection^[18]^. To increase detection of SARS-CoV-2 infection, a pooled plasma and respiratory sample test may be useful. In contrast, should clinicians like to assess the severity of the COVID-19 disease in an individual, testing SARS-CoV-2 RNA from a plasma sample alone may be beneficial. In this work, we present workflows using an in-house, inexpensive method to extract RNA from plasma, respiratory samples, or their combination.

## Materials and Methods

### Generation and characterization of in-vitro transcribed RNA standards

DNA fragments (GenBlock, Integrated DNA Technology, Coralville, IA) containing the amplification regions for HIV LTR assay (base position: 544-621 HXB2; GenBank: K03455.1) or N1/N2 assay (base position: 28,307-28,334 and 29,184-29,212 SARS-CoV-2 respectively; GenBank: NC_045512.2) with upstream T7 RNA promoter sequence was used as starting DNA construct for RNA generation. 100ng DNA construct was converted to RNA using T7 RNA polymerase (M0251; New England Biolabs (NEB), Ipswich, MA) and after digestion with DNAse I (M0303, NEB) was purified (T2040, NEB) according to manufacturer’s protocols^[21, 22]^. Product fragment lengths were confirmed using TapeStation Agilent 2200 (5067-5578), Agilent, Santa Clara, CA). Finally, Crystal digital PCR (**cdPCR**) was used to quantify the concentrations of the in-vitro transcribed RNA standard (**Supplementary Figure 1**).

### Contrived specimens for assay optimization

For HIV testing, contrived samples were prepared by adding *in-vitro* transcribed HIV RNA (for positive samples) or water (for negative samples) to negative pooled plasma specimens (IPLAWBK2E100ML, Innovative Research, Novi, MI) mixed with lysis buffer after a 10–15-minute incubation. For COVID-19 testing, negative Viral Transport Media (**VTM**) (220220, Becton Dickenson, Franklin Lakes, NJ) was spiked with 1ng/µL human genomic DNA (3041, Promega, Madison, WI), lysed in a similar manner as contrived HIV samples and then spiked with *in-vitro* transcribed SARS-CoV-2 RNA (for positive samples) or water (for negative samples).

### Panel of clinical specimens

We used a total of 113 specimens to evaluate the proposed workflows (OPTION 1 and OPTION 2, **Fig. 1a**). This remnant nasal secretion (**NS**) and plasma specimens went through several freeze-thawing cycles, and therefore were suboptimal to directly compare the original results by the certified laboratories to results obtained by the in-house workflow. Thus, in this study, we extracted the specimens using the commercial RNA extraction kit (referred as standard extraction kit) and in-house extraction kit (referred as in-house extraction kit). The extracted RNA was subsequently analyzed using the same RT-qPCR and results were compared. Discordant results were ruled out using the original results previously reported by the certified laboratories.

**Fig. 1.**
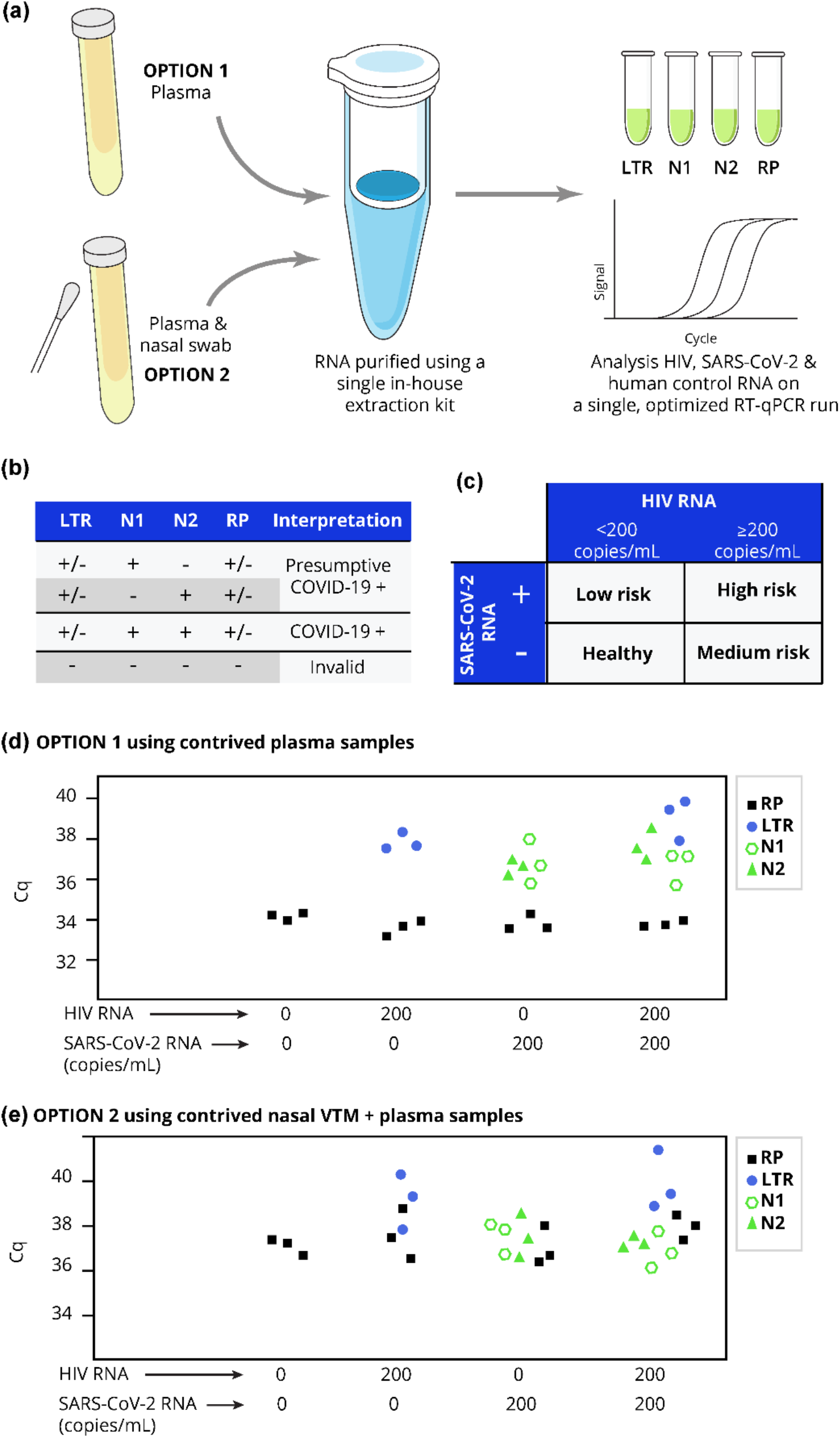
Workflows for SARS-CoV-2 screening and HIV viral load testing. (**a**) Schema of the workflows for analysis of only plasma specimens or the mixtures of plasma and respiratory specimens. Specimens are extracted using our low-cost, in-house extraction method and analyzed using RT-qPCR assays which target HIV LTR gene, SARS-CoV-2 N gene (N1, N2), and human RP gene (RP). (**b**) Assay interpretation of the results based on HIV and SARS-CoV-2 RT-qPCR results (**c**) Clinical interpretation based on the status of HIV viral load and SARS-CoV-2 infection. (**d**) Analysis of HIV and SARS-CoV-2 RNA spiked in lysed negative plasma at 0 or 200 copies/mL. (**e**) Analysis of HIV and SARS-CoV-2 RNA spiked in lysed pooled NS and plasma at 0 or 200 copies/mL. Individual Cq values of three technical replicates (individually extracted and RT-qPCR assayed) are plotted.

We calculated that we would need at least 30 SARS-CoV-2 positive specimens so that we could estimate 95%CI of 90%-100%, should all the analysis be accurate (Binomial Exact Proportion Estimation). For OPTION 1 workflow, remnant NS in **VTM** that were positive for SARS-CoV-2 (*n = 31*) and negative for SARS-CoV-2 (*n = 36*) but positive for other respiratory pathogens (*e*.*g*., influenza, seasonal coronavirus, adenovirus, parainfluenza virus, metapneumovirus, enterovirus, bocavirus, and pneumoniae were used^[23]^. These specimens were either nasopharyngeal specimens collected by health personnel or observed self-collected nasal swabs from individuals presenting respiratory symptoms, as part of the Seattle Flu Study in 2020. The swabs were suspended in 3mL VTM and previously tested in a CLIA-certified laboratory ^[24]^. Aliquots of these specimens were de-identified prior to testing in the University of Washington’s Lutz laboratory and Seattle Children’s Research Institute’s Frenkel laboratory, as part of this study.

Remnant HIV-seropositive plasma (*n = 30*) were used to evaluate the performance of the HIV viral load tests using the in-house extraction method compared to the standard extraction method. The HIV-seropositive plasma specimens were obtained from the CIENI/INER (WHO HIV certified lab) plasma bank in Mexico City in 2018, as described ^[25]^. HIV VL was measured in all the selected samples using the Abbott m2000 system (Chicago, IL) and known to contain near or above 1000 copies/mL HIV RNA. We randomly selected at least 30 specimens to allow estimation of VL. As negative control for HIV VL test, we included 10 negative plasma specimens. All samples were de-identified prior to shipment to and testing in the Seattle Children’s Research Institute’s Frenkel laboratory. We also included analysis of HIV-seronegative (*n = 10*, screened by the vendor Innovative Research, Inc, MI, USA) plasma as a negative control.

To test the OPTION 2 workflow, a subset of specimens (*n = 26*) remaining from the OPTION 1 workflow experiments were mixed with NS to create 26 HIV-seropositive specimens of which 10 were SARS-CoV-2 positive and 16 were SARS-CoV-2-negative specimens.

### Ethics statement

The remnant respiratory specimens were collected as part of Seattle Flu Study in 2020 (IRB#: STUDY0006181). Informed consent was obtained from adult participants and parents or permanent legal guardians of participant children. Archived specimens were randomly chosen for use in the current study. Participants from CIENI/INER gave written consent for the use of remnant plasma samples, approved by the Institutional Review Board of the National Institute of Respiratory Diseases (Project code: E02-17).

### In-house RNA extraction kit

Lysis buffer was freshly prepared from mixing the buffer (4M guanidine thiocyanate, 10mM MES pH 5.5) with 6% (v/v) 1mg/mL tRNA (AM7119, ThermoFisher, Waltham, MA), 2% (v/v) 2M DTT (P1771, Promega, Madison, WI). VTM (50µL), plasma (140µL), or pooled VTM (50µL)/plasma (140µL) was mixed with 4x volume of lysis buffer and incubated at room temperature for 15 minutes. The 630µL mixture was then added to the silica column (Epoch Life Sciences, 1920-250) and centrifuged at 6000g for 1 minute. Flow-through fluid was decanted, and the remaining mixture underwent a repeated centrifugation until all the lysed sample was captured by silica. After the last spin of the lysed sample, a new collection tube was placed, 800µL wash buffer 1 (1M guanidinium thiocyanate, 10mM Tris-HCl pH 7.4) was added. The tube and column were centrifuged at 6000g for 1 minute. Flow-through was discarded and a new collection tube was placed. 800µL wash buffer 2 (80% ethanol; 20% 10mM Tris-HCl pH 7.4) was added and the tube was centrifuged at 20,000g for 3 minutes. Flow-through was discarded and the tube was centrifuged at 21,000g for additional 1 minutes to completely remove any residue wash buffer. The spin column was then transferred to a clean 1.5mL tube (Lo-bind DNA, Eppendorf) and 0.04% (w/v) sodium azide in water was added to elute the RNA (60µL for either plasma or NS; and 80µL for co-extracted specimens to have sufficient samples for all the RT-qPCR experiments. After 1-min incubation at room temperature and 1-minute 6000g centrifugation, the resultant RNA was collected and stored at -80°C until tested by RT-qPCR. All samples underwent only a single freeze-thawing cycle to avoid significant RNA degradation.

### Commercial RNA extraction method (referred as standard method)

The QIAamp viral RNA kits (52904, Qiagen, Hilden, Germany) was used as a standard method to compare the performance of the in-house RNA extraction protocol using the same input volume of specimens described above. All samples were processed according to the manufacturer’s protocol^[26]^.

### RT-qPCR assays for HIV viral load and SARS-CoV-2 detection

We adopted clinically-validated primer and probe sets for detection of HIV^[31]^ and SARS-CoV-2^[32]^. We selected this LTR assay as its probe and primers have melting temperatures similar to those used in the US CDC SARS-CoV-2 N1, N2 assays. Using the same temperature cycling profile, we reliably amplified the *in-vitro* transcribed HIV and SARS-CoV-2 RNA down to 10 copies/reaction when running for 40 cycles (**Supplementary Figure 2**).

Four separate RT-qPCR reactions targeting HIV long terminal repeat (**LTR**), SARS-CoV-2 nucleocapsid gene regions N1 and N2 and human ribonuclease P (**RP**) were set up for each sample. Each 20µL RT-PCR reaction contained 1x TaqPath master mix (A15299, ThermoFisher, Waltham, MA), HIV, SARS-CoV-2 or human RP primers/probes (**Supplementary table 1**) and 5µL extracted RNA or RNA standards at 0 - 10^7^ copies/reaction. The reaction mixtures were run on a real-time thermal cycler (CFX96, Biorad) using 2 min at 25°C for primer annealing and UNG digestion; 15 min at 50°C for reverse transcription; 2 min at 95°C for reverse transcriptase enzyme deactivation and initial denaturing, and 50 cycles of 3 sec at 95°C, 30 sec at 55°C. At the end of 30 sec at 55°C, the fluorescence was read using the FAM channel. Sample copies per reaction were calculated using the RNA standard curve included in each run and used to calculate RNA copies/mL of original specimen. The number of RT-qPCR replicates are provided in the caption of each experiment. RT-qPCR data were analyzed using CFX Maestro (Biorad). Data were normalized using cycles 5-10; and samples were determined positive when the normalized signals were above 200 RFU. The Cq values of samples were interpolated based on the linear fit to the Cq values and log10 of input concentration of the RNA standards from 0-10^7^ copies/reaction.

### Interpretation of measured VL results and statistical analysis

In contrived specimens, the measured VL from RNA extracted by both methods were plot against the spiked in RNA input concentration to demonstrate the performance of each extraction method. The measured VL were classified as “correct” if they fell within the upper and lower bounds of precise measurements defined as ±0.3log10(RNA copies/mL)^[27]^. For HIV-seropositive specimens, measured VL values from the RNA extracted by in-house vs standard methods were assessed for their correlation. R^2^ values from a linear regression fit of scattered plots and p-values from Wilcoxon Signed-Rank test (two-tailed) were reported. Next, we used the US NIH VF cutoff^[28]^ or the WHO VF cutoff ^[29]^ to classify specimens as VL suppressed “**VLS**” (VL<200 or VL<1000 copies/mL, respectively) or virologic failure “**VF**” (VL≥200 or VL≥1000 copies/mL, respectively). % Concordance of the HIV VL classifications from RNA extracted by the in-house vs the standard RNA extraction method was calculated from number of concordant VL classifications (i.e., VLS vs VF)/ number of seropositive samples tested. Similarly, for SARS-CoV-2 specimens, measured VL values from the RNA extracted by in-house vs standard methods were assessed for the correlation using a linear regression fit and for its statistical difference using Wilcoxon Signed-Rank test (two-tailed). Next, we classified results as “negative,” “positive,” or “presumptive positives,” based on the results of N1, N2, and RP assays. Positive samples have detection of N1 and N2 regardless of the results from the RP assay. Presumptive samples have detection of either N1 or N2 regardless of the results from the RP assay. Negative samples are negative for both N1 and N2 but are positive for RP. If samples are not detected of N1, N2, and RP, they are classified as indeterminate results. % Concordance of the SARS-CoV-2 results by the in-house vs standard RNA extraction method was calculated from the # of samples with concordant results (i.e., negative, positive, presumptive positive, and indeterminate)/ # of samples tested.

## Results

### Integrated workflow for HIV viral load test and SARS-CoV-2 screening

SARS-CoV-2 RNA has been detected in both plasma and respiratory specimens; and thus, we have envisioned two options for screening SARS-CoV-2 infection while performing routine HIV viral load tests (**Fig. 1a**). OPTION 1 utilizes plasma specimens as the sole sample source for HIV RNA load test and SARS-CoV-2 screening. The extracted RNA is subjected to four RT-qPCR reactions; two non-overlapping regions in the SARS-CoV-2 nucleocapsid gene (**N1** and **N2**), HIV long terminal repeat (**LTR**) gene, and human ribonuclease P (**RP**) gene. If samples test negative for all assays, this suggests procedural errors, and re-testing and/or re-collecting samples is recommended (**Fig. 1b**). RT-qPCR results report HIV VL and SARS-CoV-2 infection status (**Fig. 1c**), individuals present different levels of risks to develop poor health outcomes. Because SARS-CoV-2 viral loads in respiratory specimens are generally higher than in plasma, we propose an alternative workflow (OPTION 2) to increase the sensitivity of SARS-CoV-2 detection, by combining the extraction of plasma/NS for detection. RNA extraction in OPTION 1 and 2 workflow is performed using an in-house Boom’s method. Analysis of contrived plasma and contrived pooled plasma/NS samples confirmed detection of 200 copies/mL of HIV and SARS-CoV-2 RNA (**Fig. 1, d** and **e)**. Based on the definition of virologic failure (**VF**) by the US National Institutes of Health^[28]^, this limit of detection (**LoD**) of HIV assay at 200 copies/mL would allow differentiation of samples collected from PLH with VF (>200 copies/mL, as these would be detected) versus viral load suppressed (**VLS**, undetected). For the SARS-CoV-2 RNA, the 200 copies/mL is lower than the reported LoD of the US CDC SARS-CoV-2 assay^[30]^. In all contrived samples, we appropriately detected the human RP gene control (**Fig. 1e**).

### Development of RNA extraction protocol

The in-house RNA extraction protocol is based on the Boom’s method,^[33]^ wherein RNA is bound to a silica column, followed by washing steps to remove PCR inhibitors and then eluted out in a low salt buffer. Herein, a lysis buffer comprised of 4M guanidinium thiocyanate (**GuSCN**), a potent chaotropic salt, was used to lyse virus and deactivate RNases, as well as serve as a salt bridge between negatively-charged RNA and negatively-charged silica, although 2M GuSCN was previously shown to be sufficient^[34]^. In addition to GuSCN in the lysis buffer, we used a combination of a reducing agent (Dithiothreitol, DTT) to deactivate RNases via reduction of disulfide bonds and sacrificial non-HIV RNA molecules (tRNA) in a relatively low pH buffer (MES, pH 5). RNA phosphodiester bonds are less prone to hydrolysis at pH 4-5^[35]^. tRNA and DTT are prone to oxidation and unstable in the liquid form at room temperature, so we stored them at -20 ºC and mixed them with GuSCN and MES lysis buffer just before the extraction experiments. A more stable reducing agent, β mercaptoethanol, has been successfully used in lysis buffer for RNA extraction^[36]^ but DTT is less toxic^[37]^ and less pungent than β mercaptoethanol. The optimal concentration of 20-40mM DTT (**Supplementary figure 3**) showed 30% recovery of spiked HIV RNA compared to the standard RNA extraction kit. We hypothesized that the rate of viral lysis may be faster than the rate of RNase deactivation, thus allowing RNases to remain active and digest HIV RNA released from the viral capsid. Addition of tRNA as sacrificial molecules with DTT improved the yield of HIV RNA. We found amplifiable HIV RNA yields were comparable to the standard RNA extraction kit when 22.4-33.6µg non-target tRNA was added to the lysis mixture (**Supplementary figure 4**). It is important to note that addition of tRNA beyond 6µg in the lysis buffer did not add additional protection to HIV RNA. The silica column we used (**Supplementary table 2**) has a high capacity to capture up to 40µg and should accommodate absorption of both tRNA and HIV RNA targets, but the tRNA eluted along with the target HIV RNA could compete during the RT-PCR amplification step. (**Supplementary figure 5**). In fact, we observed inefficient target amplification when non-target DNA is present at more than 2 ng/µL depending on the PCR reaction conditions^[38, 39]^. To wash loosely-bound charged molecules from the silica membrane and further purify the RNA, we used two wash buffers starting from a high salt wash buffer: 1M GuSCN, 10mM Tris-HCl pH 7.4; and followed by a low-salt wash buffer: 10mM Tris-HCl pH 7.4 with 80% ethanol. A high concentration of ethanol can solubilize residual salts but not RNA from the silica membrane when GuSCN is absent to serve as a salt bridge.

### Analytical sensitivity in contrived specimens

The analytical sensitivity of OPTION 1 and OPTION 2 workflows using in-house extraction method was assessed and directly compared to the standard extraction method. For the OPTION 1 workflow, we analyzed single-target contrived samples (*i*.*e*., either HIV or SARS-CoV-2 RNA spiked in lysed human plasma). **Fig. 2a** and **Fig. 2b** show scatter plots of the input VL spiked in contrived samples and measured VL from the RNA extracted by in-house or standard methods. Across the concentrations from the LoD to 60K copies/mL, the in-house extraction method has a 1.09 ± 0.5 (mean ± SD) efficiency (slopes of fitted linear regression, **Fig. 2a** - **2b**), compared to 0.94 ± 0.1 efficiency of the standard method. Regardless of the extraction methods used, classifications as positive (detectable VL) and negative (undetectable VL) revealed RNA from the in-house extracted method was successfully detected in 100% (12/12) positive samples spiked with ≥200 copies/mL HIV RNA and undetected in 3/3 negative samples. Analysis of RNA from contrived SARS-CoV-2 samples, using in-house versus standard extraction methods, showed 100% agreement for the positive (detectable VL) and negative (undetectable VL) classifications (**Fig. 2b**), with an LoD of 100 copies/mL in positive samples. For the OPTION 2 workflow (pooled plasma/NS), the in-house extraction method yielded LoDs identical to those obtained from the OPTION 1 workflow (*i*.*e*., 200 copies/mL for HIV; 100 copies/mL for SARS-CoV-2, **Supplementary figure 6**). HIV VL measured in the RNA obtained from both extraction methods are in close agreement (linear regression fit R^2^: 0.98 for measured VL correlation between the two different extraction methods from plasma (y-axes in both plots in **Fig. 2a**); 0.92 for measured VL correlation between the two extraction methods from pooled plasma/NS samples (y-axes in both plots in **Supplementary figure 6a**). The yields of HIV RNA from both extraction methods were found to be more variable in ≤200copies/mL samples (**Fig. 2a** – **2b**, black diamonds are samples with measured VL significantly deviated from the theoretical yield). This observation is expected because the theoretical RNA concentration per RT-qPCR reaction is very low (i.e., <7 copies/reaction for the starting 140µL of samples with <600 copies/mL). The variations of measured VL in <600copies/mL samples are also observed in pooled plasma/NS (**Supplementary figure 6**). We used a 50-cycle RT-qPCR (while 40-cycle RT-qPCR is common) to increase the opportunity to detect low-concentration RNA in 100-200 copies/mL samples. However, the limit of quantification (**LOQ**) of HIV VL in contrived plasma or plasma/NS is 600 copies/mL, higher than the LoD. **Fig. 2c** and **Fig. 2d** summarize the performance of both OPTION 1 and OPTION 2 workflows on SARS-CoV-2 RNA detection (*i*.*e*., positive vs negative) and HIV VL classifications (*i*.*e*., VLS vs VD) based on the US NIH VF threshold.

**Fig. 2.**
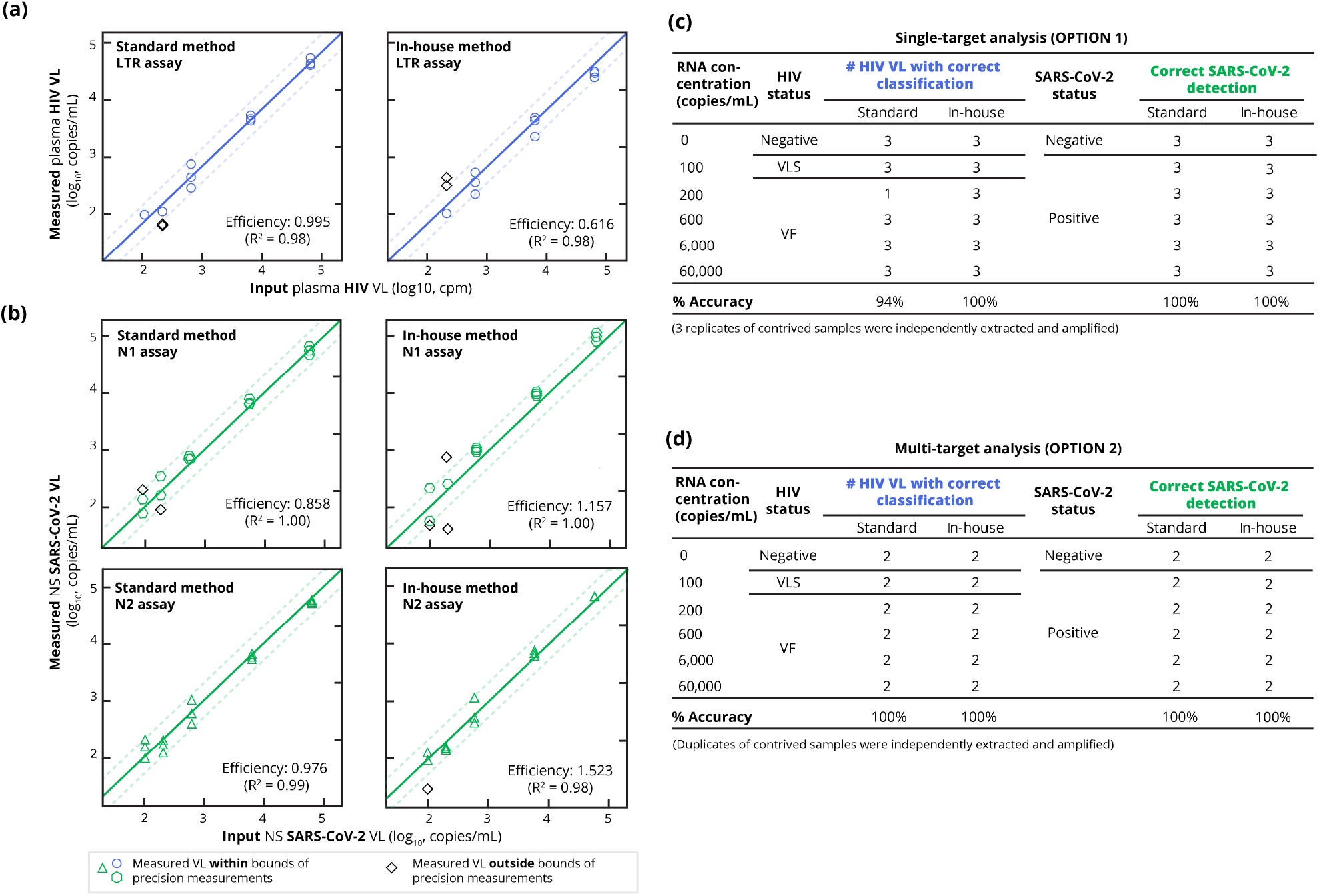
Quantitation and classification of HIV and SARS-CoV-2 RNA obtained from in-house and standard extraction methods. Scatter plots of (**a**) measured HIV VL (*n = 3*) vs input RNA in contrived plasma samples (lysed plasma and spiked with synthetic HIV RNA at 0, 100, 200, 600, 6K, or 60K copies/mL), and (**b**) measured SARS-Cov-2 VL (*n = 3*) vs input RNA in contrived NS (lysed and spiked with synthetic SARS-CoV-2 RNA at 0, 100, 200, 600, 6K, or 60K copies/mL). Diagonal lines represent 100% theoretical RNA recovery from the extraction method based on the spiked-in RNA input. Dashed diagonal lines indicate the bound for precise measurement (*i*.*e*., measured VL within ±0.3 log_10_(VL, copies/mL of input)). Efficiency of each extraction method for each assay is the slope of the linear regression fit, reported along with its R^2^. Summary of HIV VL classification and SARS-CoV-2 detection using both extraction methods using (**c**) OPTION 1 workflow to analyze either NS or plasma, and (**d**) OPTION 2 workflow to analyze pooled plasma/NS samples. For HIV samples, we classified results as VLS or VF. For SARS-CoV-2, we classified results as positive or negative based on whether they are detected or not detected by both N1 and N2 assays. **Supplementary figure 6** for the scatter plots of the OPTION 2 workflow. Data in **Fig. 2** and **Supplementary figure 2** were collected by a single assay operator.

### Clinical evaluation of HIV in plasma specimens

**Fig. 3a** shows our experimental design to compare the performance of standard versus in-house extraction methods. RNA extracted using the in-house method (**Fig. 3b**) was detected in 100% (30/30; 95%CI: 88-100) of HIV-seropositive plasma (median VL: 30,604 copies/mL; IQR: 6,503 - 96,626 copies/mL) and not falsely detected in HIV-seronegative plasma (10/10). Human RP genes were detectable in all negative specimens, confirming a functioning assay. However, human RP levels in HIV-seronegative plasma were significantly higher (lower Cq values) than those of HIV-seropositive plasma (**Fig. 3b** and **Supplementary figure 7**). Measured HIV VL levels in the RNA extracted by in-house and standard methods were positively correlated (**Fig 3c**, linear regression fit, R^2^: 0.81) but were statistically different (p<0.05, Wilcoxon Signed-Rank test, two-tailed). Compared to the standard method (**Fig. 3c**), 20/30 measured VL values of RNA extracted by the in-house method were within the precise measurement bound, but RNA extracted by the in-house method led to 7 overestimated VL values and 3 underestimated VL values (black diamonds). Regardless, classifications (*i*.*e*., VLS vs VF) of measured VL results (**Fig. 3d**) from RNA obtained by both extraction methods, either at ≥200 copies/mL for VF threshold (US NIH guideline^[28]^) or ≥1000copies/mL for VF (WHO guideline^[29]^), had 100% concordance (30/30; 95% CI: 90-100).

**Fig. 3.**
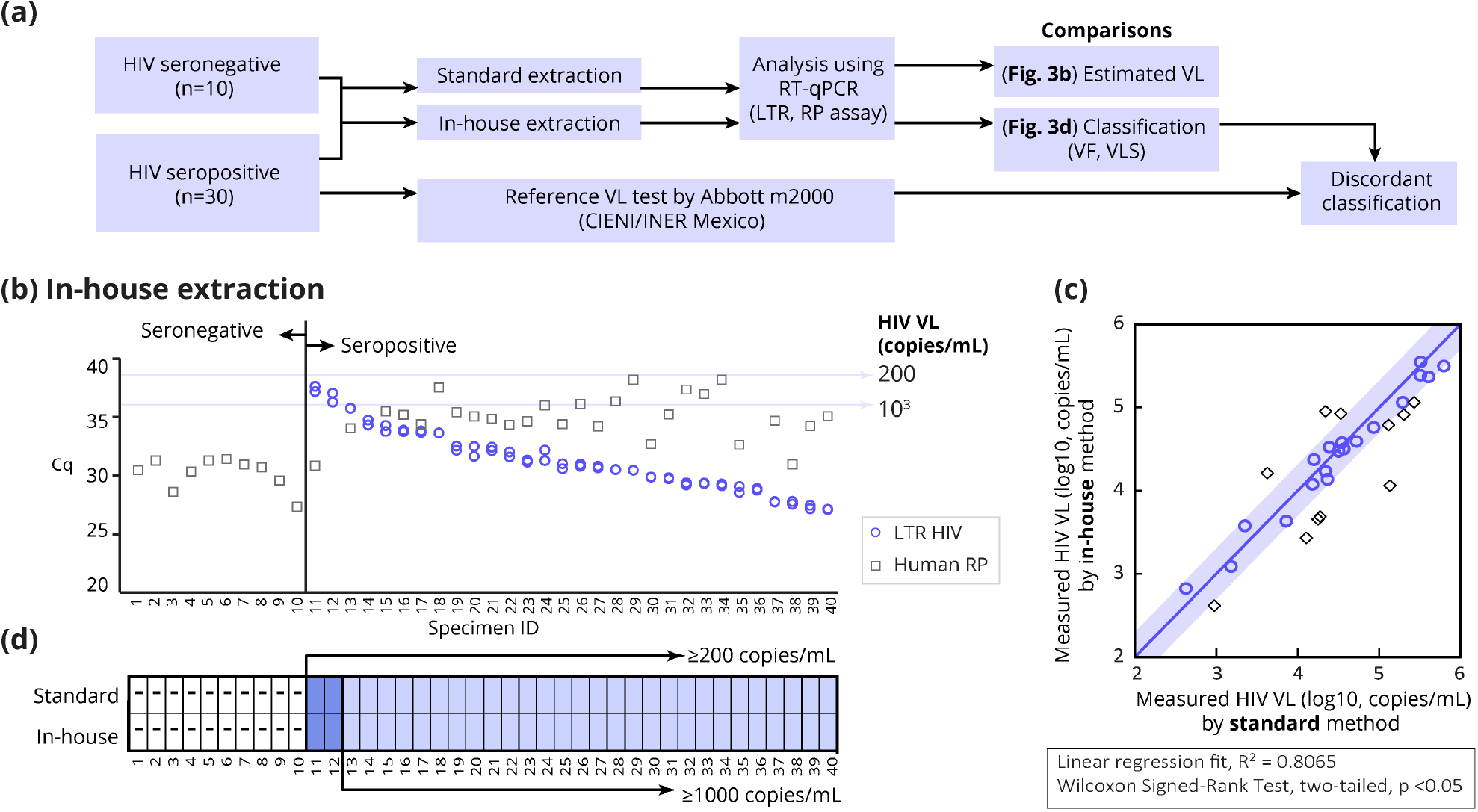
Quantitation and classification of HIV RNA in clinical plasma specimens. (**a**) Schema of experimental design to compare the performance of standard vs in-house extraction methods. Plasma RNA extracted by standard or in-house extraction was evaluated using RT-qPCR. The RT-qPCR results were compared in terms of measured viral load and classifications. All HIV seropositive specimens had VL ≥1000 copies/mL when tested by the reference lab using Abbott m2000. (**b**) Cq values of the HIV long-terminal repeat (LTR) assay and human ribonuclease P (RP) assay (control for negative specimens) from the plasma RNA extracted by the in-house method. Extracted RNA from HIV-seropositive specimens were analyzed by LTR and RP RT-qPCR assays in duplicate. HIV-seronegative specimens (as tested by the vendor) were each extracted twice, and each extracted RNA aliquot was analyzed by LTR and RP RT-qPCR assays. The average RP Cq values are plotted. (**c**) Scatter plot of measured HIV VL in specimens extracted by standard vs in-house methods. The diagonal line represents a theoretical 100% correlation of HIV VL measured in RNA extracted from both extraction methods. The shaded box around the diagonal lines indicate precise measurement bound (±0.3 log_10_(VL measured in RNA extracted by standard method, copies/mL)). Diamonds are samples with measured VL outside precise measurement bound. (**d**) Classifications correspond to VL results. Negative results are labeled as “-” in white boxes. Virologic failure results were classified using 200 copies/mL (dark purple boxes) and 1000 copies/mL (light purple boxes) thresholds. RNA extracted by both methods was collected by one assay operator and analyzed by RT-qPCR by another assay operator.

### Clinical evaluation of SARS-CoV-2 in NS specimens

**Fig. 4a** shows our experimental design to compare the performance of standard versus in-house extraction methods. NS was previously tested in a CLIA-certified laboratory, as 31 positive and 36 negative NS specimens for SARS-CoV-2. In this study, due to limited volume of specimens available, we extracted 50µL specimens instead of 140µL (as was done in the above contrived sample experiment). Of 31 specimens previously-tested positive for SARS-CoV-2 (**Fig. 4b** and **Fig. 4c**), the in-house method detected 25 (81%) as positive (detected by the N1 and N2 assay; median VL: 4180 copies/mL; IQR:3912-176168 copies/mL) and 6 (19%) as presumptive positive (detected by either the N1 or N2 assay; median VL: 84 copies/mL; IQR: 39–203 copies/mL). Two SARS-CoV-2 specimens (VL: 162 and 268 copies/mL) detected by the in-house method (confirmed true positive by the certified lab results) were falsely negative and presumptive positive by the standard method. Measured concentrations of the SARS-CoV-2 RNA obtained by in-house versus standard extraction methods revealed high correlations (R^2^: 0.95 for N1 in **Fig. 4d**; 0.99 for N2 in **Fig. 4e**) and had no significant difference (p>0.05; Wilcoxon Signed-Rank test, two-tailed). Comparing the measured VL from the RNA obtained from the in-house and standard extraction methods, we found a few measured VL values in N1 assay outside the precise measurement bound (**Fig. 4d**), but the measured VL values from the same specimens were within acceptable variation in N2 assay (**Fig. 4e**). Possibly, this is due to the reportedly lower sensitivity of the N1 assay^[32]^, thus the N1 assay could not detect low concentrations of the SARS-CoV-2 RNA in these specimens (VL: <500 copies/mL from the 50µL sample, equivalent to <178 copies/mL from a 140µL sample, which is around the LoD we observed from the contrived specimens). Similarly, the RP assay had 4 and 2 indeterminate (**IND**) results in the workflow using RNA obtained from the in-house and standard methods, respectively. These specimens contained <250 copies/mL human RP, thus, this discordance is likely due to variations of RT-qPCR near the LoD. **Fig. 4f** summarizes the SARS-CoV-2 detection results from the RNA obtained by standard and in-house extraction methods.

**Fig. 4.**
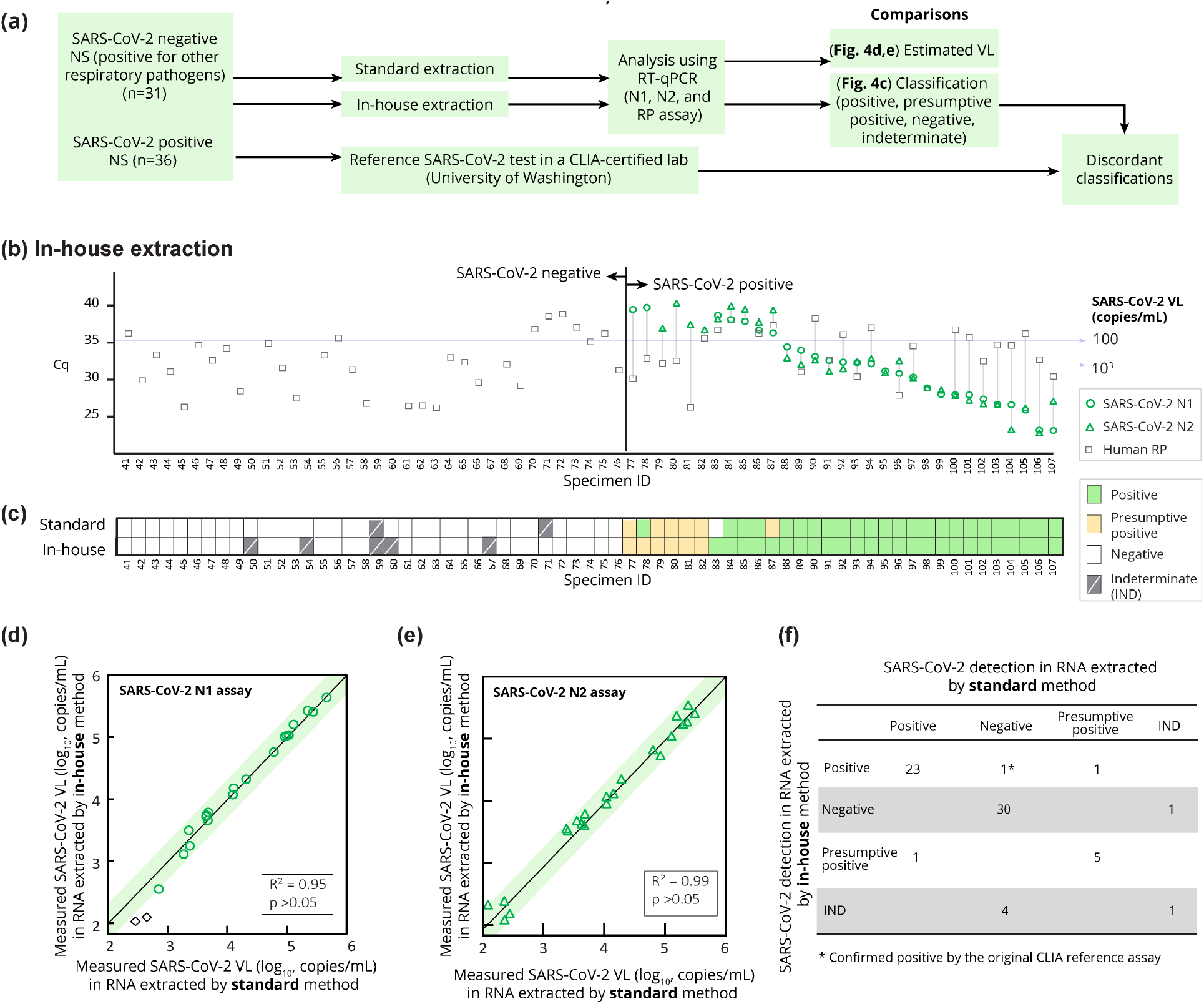
Quantification and detection of SARS-CoV-2 RNA in clinical NS specimens. (**a**) Schema of experimental design to compare the performance of standard vs in-house extraction methods. NS RNA obtained from standard or in-house extraction were evaluated using RT-qPCR. The RT-qPCR results were compared in terms of measured viral load and classifications. Discordant classifications were ruled out based on previous results from the CLIA-certified lab. (**b**) Cq values of the SARS-CoV-2 N1, N2, and human RP assay (control for negative specimens) from the in-house method. Each specimen was extracted, followed by RT-qPCR, and represented as individual data points. (**c**) Classifications of specimens based on the results from the two extraction methods. Specimens detected by N1 and N2 (regardless of RP detection) are classified as positive; specimens detected by either N1 or N2 assays are classified as presumptive positive; specimens not detected by N1 and N2 but detected by RP assay are classified as negative; specimens not detected by any assays are classified as indeterminate (**IND**). Scatter plot of measured SARS-CoV-2 in specimens extracted by standard vs in-house methods in (**d**) N1 assay (**e**) N2 assay. The diagonal lines represent a theoretical 100% correlation of SARS-CoV-2 VL measured in RNA extracted from both extraction methods. Dashed lines indicate RT-qPCR precise measurement bound (*i*.*e*., ±0.3log10(VL measured in RNA extracted from standard method, copies/mL). Diamonds are samples with measured VL that significantly deviates from those obtained via standard extraction. (**f**) Summary table for classifications by the two methods. Data were collected by two assay operators.

### Clinical evaluation of HIV and SARS-CoV-2 in pooled clinical plasma and NS

We tested the OPTION 2 co-extraction workflow on a subset of available remnant NS and plasma specimens (**Fig. 5a**) previously tested in the CLIA-certified and reference labs, respectively. Here, each HIV-seropositive plasma specimen (*n = 26*) was randomly assigned to mix with either positive (*n = 10*) or negative SARS-CoV-2 specimens (*n = 16*). Each plasma/nasal specimen pool was then extracted using the in-house method and the standard method. **Fig. 5b** shows the Cq values of SARS-CoV-2 N1 and N2, HIV LTR, and human RP assays from the samples extracted by the in-house method. Using the US NIH 200copies/mL cutoff for HIV VF (**Fig. 5c**), there was a 100% (26/26) concordance with the standard method. Detection of SARS-CoV-2 was also 100% (26/26) concordant. **Fig. 5d** – **Fig. 5f** show high correlations of measured VL of HIV LTR, SARS-CoV-2 N1, and SARS-CoV-2 N2 targets, respectively. Interesting, the measured HIV VL values in these specimens from RNA obtained by the in-house method were significantly higher than those by the standard method (p<0.05, Wilcoxon Signed-Ranked test, two-tailed); but the measured VL of SARS-CoV-2 are similar between the two groups.

**Fig. 5.**
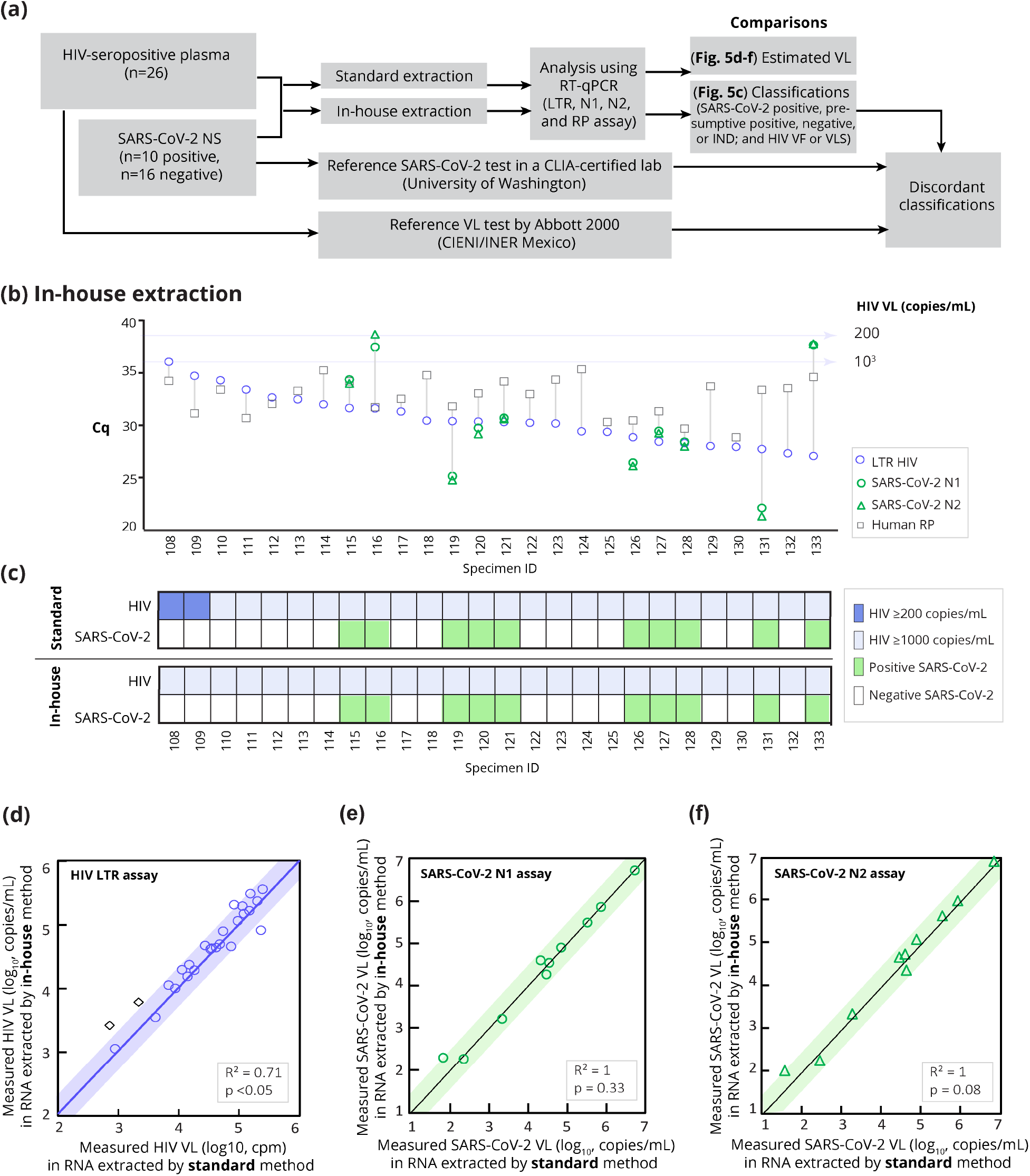
Comparison of HIV LTR and SARS-CoV-2 Cq values obtained from RNA co-extracted from NS/plasma specimens using in-house vs standard extraction protocols. (**a**) Schema of experimental design to compare the performance of standard vs in-house extraction methods. NS/plasma RNA extracted by standard or in-house extraction were evaluated using RT-qPCR. The RT-qPCR results were compared in terms of measured VL and classifications. (**b**) Cq values of the SARS-CoV-2 LTR, N1, N2, and human RP assay (control for negative specimens) from the in-house method. Each specimen was extracted, followed by RT-qPCR, and represented as individual data points. (**c**) Classifications of specimens based on the results from the two extraction methods. Scatter plot of measured SARS-CoV-2 in specimens extracted by standard vs in-house methods in (**d**) LTR (**e**) N1 (**f**) N2 assay. The diagonal lines represent a theoretical 100% correlation of VL measured in RNA extracted from both methods. Dashed lines indicate precise measurement bound. Diamonds are samples with measured VL that significantly deviates from those obtained via standard extraction.

## Discussion

We developed a new workflow with an optimized in-house RNA extraction method to enable simultaneous quantification of HIV VL and detection of SARS-CoV-2 in a single RT-qPCR run. The workflow can be tailored to different clinical use by isolating target RNA from plasma or plasma pooled with NS via a single extraction kit.

In contrived specimens, both OPTION 1 and OPTION 2 workflows can detect low concentrations of the HIV RNA (200 copies/mL) and SARS-CoV-2 RNA (100 copies/mL). We anticipate that this highly sensitive SARS-CoV-2 assay would be able to detect most SARS-CoV-2 positive clinical specimens, as the LoD of 100 copies/mL is lower than the reported medians of SARS-CoV-2 VL in blood in the first two weeks and ∼50-fold lower than the medians of SARS-CoV-2 viral loads in respiratory samples during the first 6 weeks^[18]^. When testing clinical NS, we found that the in-house method had positive results for all SARS-CoV-2 respiratory specimens with ≥1000 copies/mL but only around 50% positive and 50% presumptive positive results in 100-1000 copies/mL specimens. Due to limited availability of specimens, we only used 50µL specimens from 1mL swab eluate. Processing a larger volume of specimens (i.e., 140µL per extraction like the case of HIV specimens or contrived SARS-CoV-2 samples), would likely improve detection of specimens with <1000 copies/mL by increasing the RNA input in the final RT-PCR reaction. Measured HIV VL in contrived specimens by both OPTION 1 and OPTION 2 also showed near 100% correlations. In clinical plasma specimens, the in-house method also accurately classified HIV VF compared to the standard method. The new workflow can potentially streamline HIV management and SARS-CoV-2 detection by determining if a PLH has detectable viremia as well as SARS-CoV-2 infection by extracting RNA from plasma alone or pooled plasma/NS using a single in-house method. Combining the detection of these two viruses can help address the challenges associated with the overlay of the COVID-19 pandemic on the existing HIV epidemic.

Our workflow utilizes an internal control based on detection of the human RP gene, which is also used for many clinical assays^[40-42]^. In addition to reporting proper assay function, detection of the RP gene can confirm whether the samples were properly collected especially for the self-collection of NS for SARS-CoV-2. As an alternative to human genes, MS2 bacteriophage can be spiked into the sample as the control^[47, 48]^. For this approach, the level of MS2 will not reflect proper sample collection or release. Rather, the MS2 bacteriophage can be used as an engineering tool to accurately measure how well the extraction and amplification are performed.

The workflow also helps to circumvent the issues associated with the availability of commercial RNA extraction kits and reduces the cost of RT-qPCR. Because of a high demand for COVID-19 tests, procuring of RNA extraction kits has been difficult, which also limits routine HIV testing. RNA extraction kits have contributed significant reagent cost for most RT-qPCR workflows (approximately $6/kit). Our in-house method uses low-cost alternatives that can be obtained as off-the-shelf reagents (**Supplementary table 2**), which allows laboratories to assemble their own low-cost RNA extraction kits ($1.85/kit). Using the proposed workflow to analyze the plasma/NS with a single in-house kit would require $1.85, 5-times less expensive for consumables than a traditional workflow requiring two commercial kits ($12) for each specimen.

Our workflow detects HIV, SARS-CoV-2, and human RNA via separate RT-qPCR reactions to maximize sensitivity. Co-amplification of SARS-CoV-2 and human RNA was performed but reportedly had reduced sensitivity compared to single-target amplification^[43]^. To improve the detection of the co-amplification, sequence-specific extraction of RNA from HIV or SARS-CoV-2 ^[38, 44-46]^ could be useful by increasing the target concentrations. Although the cost of single-target RT-PCR amplification using FAM can be expensive, it is possible to eliminate the need for the thermal cycler. End-point analysis of single channel fluorescence such as FAM can be analyzed via cell phone images and has been shown to have a high correlation with the Cq values^[23]^. Using multiplexed amplification may require a higher cost RT-qPCR than a $5000 single-channel qPCR machine, but the overall cost may be offset by higher throughput, but could reduce per-test cost since one sample can be analyzed in a single RT-qPCR reaction per sample instead of four RT-qPCR reactions per sample.

Overall, utility of such effective workflows for PLH is not limited to SARS-CoV-2 but should be explored for other common HIV comorbidities (*e*.*g*., tuberculosis, hepatitis B, hepatitis C), or other AIDS-defining opportunistic infections still causing high disease burden in low and middle income countries (*e*.*g*., *Pneumocystis jirovecii* pneumonia, histoplasmosis, toxoplasmosis, cryptococcal meningitis and mycobacteriosis). This allows the pre-scheduled VL tests to screen for other diseases that could worsen the health of PLH, reducing diagnosis costs and time.

## Data Availability

All data were available in main text or supplementary information.

## Acknowledgments

We thank Dr. Lucia Vojtech for training and access to the equipment needed for the cdPCR experiment. The Naica cdPCR system from Stilla Inc. was funded by the NIH-funded Centers for AIDS Research (P30 AI027757 and P30 AI064518). We thank other Lutz Lab members: Ian Hull, Qin Wang, and Jordan Campbell for their technical support. We thank the Seattle Flu Study and the Seattle Coronavirus Assessment Network (SCAN) teams led by Principal Investigators: Helen Y. Chu, MD, MPH, Michael Boeckh, MD, PhD, Janet A. Englund, MD, Michael Famulare, PhD, Barry R. Lutz, PhD, Deborah A. Nickerson, PhD, Mark J. Rieder, PhD, Lea M. Starita, PhD, Matthew Thompson, MD, MPH, DPhil, Jay Shendure, MD, PhD and Trevor Bedford, PhD for providing specimens for testing.

## Author contributions

NP envisioned the integrated workflow for simultaneous HIV VL testing and COVID-19 screening GG, NP, SWAS, AKO, IB, and CB conducted the experiments. GG and NP performed data analysis. NP led data visualization. SA, PDH, GRT, and LS characterized and provided specimens used in this evaluation. IB and LF provided technical insights on discussion. BRL and JJL oversaw the study. All authors contributed to writing this manuscript and approved the final version for submission.

## Supplementary Information list

**Supplementary figure 1**. Characterization of in vitro transcribed RNA standards using Crystal digital PCR (cdPCR).

**Supplementary figure 2**. Standard curves of RT-qPCR assays

**Supplementary figure 3**. Effect of DTT concentrations in in-house lysis buffer (4M GuSCN, 10mM MES pH 5.5)

**Supplementary figure 4**. Effect of carrier RNA in in-house lysis buffer (4M GuSCN, 10mM MES pH 5.5, 4% DTT).

**Supplementary figure 5**. Impact of carrier tRNA concentration on RT-qPCR

## Supplementary information

**Supplementary figure 1.**
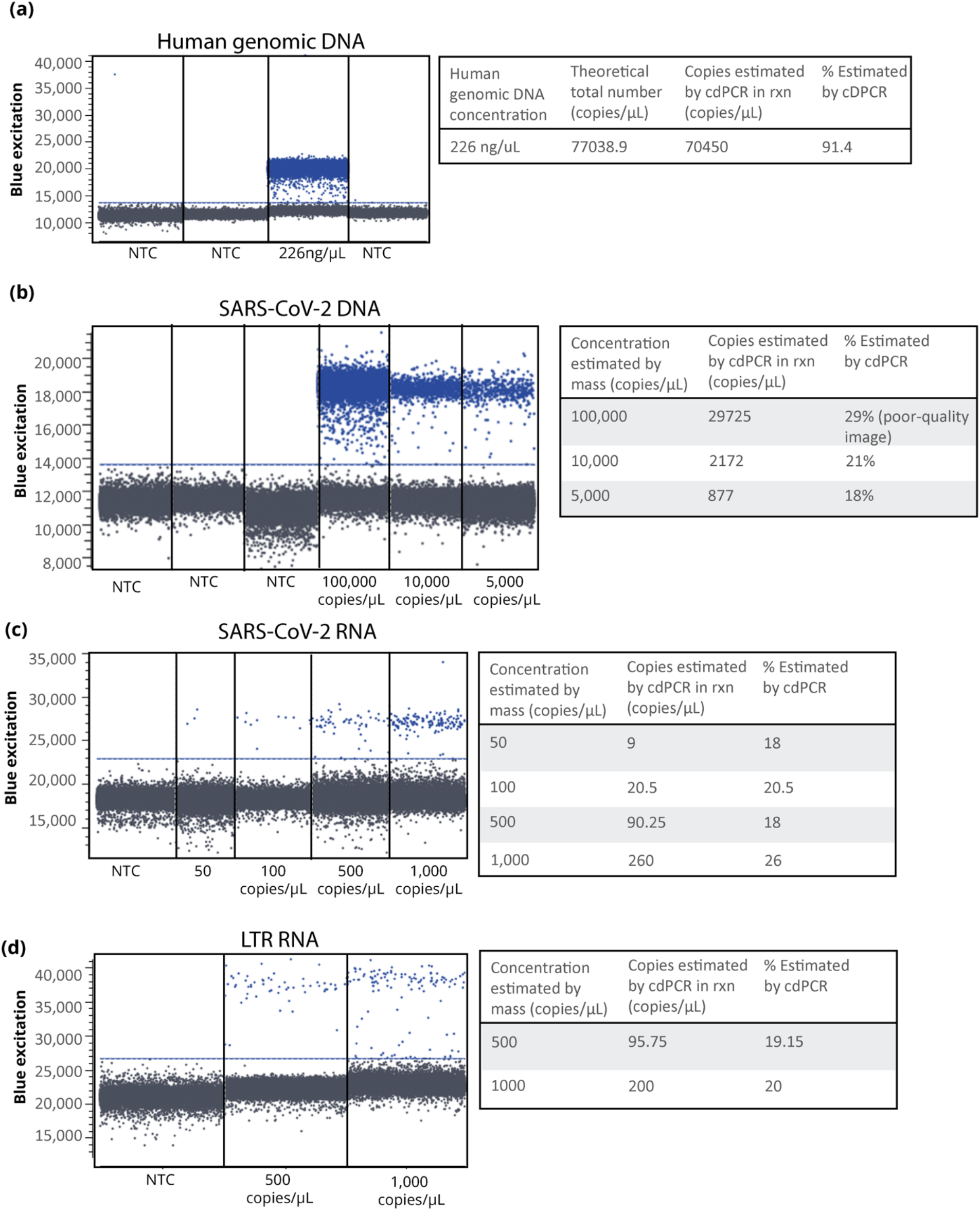
Characterization of in vitro transcribed RNA standards using Crystal digital PCR (cdPCR). (**a**) Quantification of human genomic DNA with known concentration (226ng/µl, Promega) shows accurate estimation of concentration by cdPCR (**b**) Estimation of HIV RNA concentration by cdPCR shows ∼20% of concentration measured by mass using Nanodrop instrument (ThermoFisher Scientific, US) (**c**) The measured concentrations based on the total mass of in vitro transcribed HIV RNA and COVID RNA was verified using cdPCR. We found that estimation of COVID RNA concentrations by cdPCR using N2 assay shows ∼20% of concentration and thus we have corrected the concentration of the RNA stocks based on these cdPCR. The Naica Geode (cdPCR) was programmed to perform PCR thermal cycling program: 95 °C for 10minutes, followed by 55 cycles of 95 °C for 30 seconds and 60 °C for 30 seconds. Image acquisition was performed using the Naica Prism3 reader in the blue channel (excitation range 415-480 nm and emission range 495-520 nm) at exposure time of 100 ms. Droplet counts were enabled by the detection of reference dye FITC in the blue channel and was performed by the Crystal Reader software. Extracted fluorescence value for each droplet was analyzed by Crystal Miner software. Blue threshold line was adjusted manually above the negative droplets of NTC sample in the Crystal Miner software.

RT-qPCR optimization was performed to reliably detect 10 copies of RNA template on HIV LTR (**Supplementary figure 2a**) and SARS-CoV-2 N1 (**Supplementary figure 2b**) & N2 (**Supplementary figure 2c**) assays.

**Supplementary figure 2.**
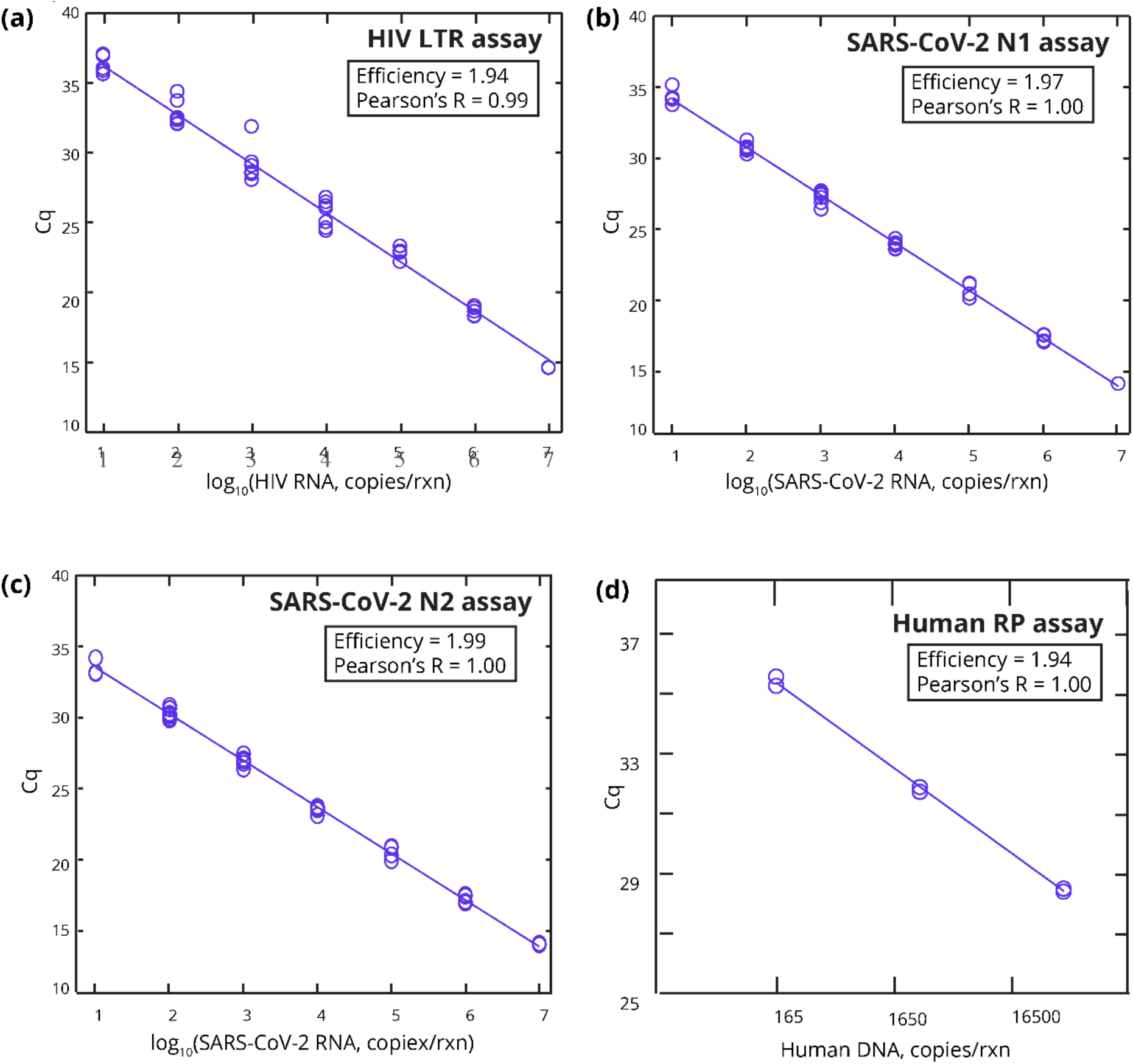
Standard curves of RT-qPCR assays. (**a**) HIV LTR assay (**b**) SARS-CoV-2 N1 assay (**c**) SARS-CoV-2 N2 (**d**) human genomic DNA For (**a**), (**b**), and (**c**), single-use stock of the standard RNA templates quantified by cdPCR (**Supplementary figure 1**) were diluted in nuclease-free water down to 2 - 2,000,000 copies/uL. 5µL of each RNA standard or water was added to 15µL of the qPCR master mix and subjected to RT-qPCR cycling as described in the methods section. Two replicate reactions were performed on each plate and data were pooled from four experiments on different days, accounting for a total of eight replicates. For human controls (**d**), two replicates/concentration were run in RT-qPCR. The concentrations were based off the reported values from the manufacturer (Promega). Note that to allow comparison across plates, we normalized the data of cycles 5-10 and used a fixed threshold of 200 RFU to determine the Cq values. All replicates were plotted along with the fitted linear regression. Corresponding Pearson’s R values were reported along with the RT-PCR efficiency (10^−1/slope of fitted linear regression line^).

### Optimizing the lysis buffer with DTT and tRNA using HIV LTR RT-qPCR assay

To optimize the lysis buffer for RNA extraction, variable DTT concentrations (2-8%) were tested. The LTR RNA spiked plasma samples were mixed with lysis buffer (added 2M DTT, 2-8% v/v) and extracted with in-house RNA extraction kit. The extracted RNA recovery was measured by RT-qPCR assay for LTR gene. **Supplementary figure 3** showed the effect of 2%, 4% and 8% (v/v) DTT in the lysis buffer on the extraction recovery of RNA as compared to standard method. 2% to 4% (v/v) of 2M DTT added in the in-house lysis buffer improved the extraction recovery of RNA copies as detected by RT-qPCR. However, recovery of RNA copies was significantly less (∼30%) as compared to standard method. We hypothesized that this reduced recovery was due to uninhibited RNAse activity in the samples. To further improve the RNA extraction recovery, tRNA was added in the lysis buffer with 4% v/v (2M) DTT. **Supplementary figure 4** showed measured HIV RNA input copies against actual HIV copies spiked in lysed plasma. Recovered HIV copies in 4-6% v/v yeast tRNA groups were comparable to standard method (1% tRNA in lysis buffer, Qiagen RNA extraction kit). This suggested that in-house lysis buffer can be used as an alternative extraction buffer for HIV RNA.

**Supplementary figure 3.**
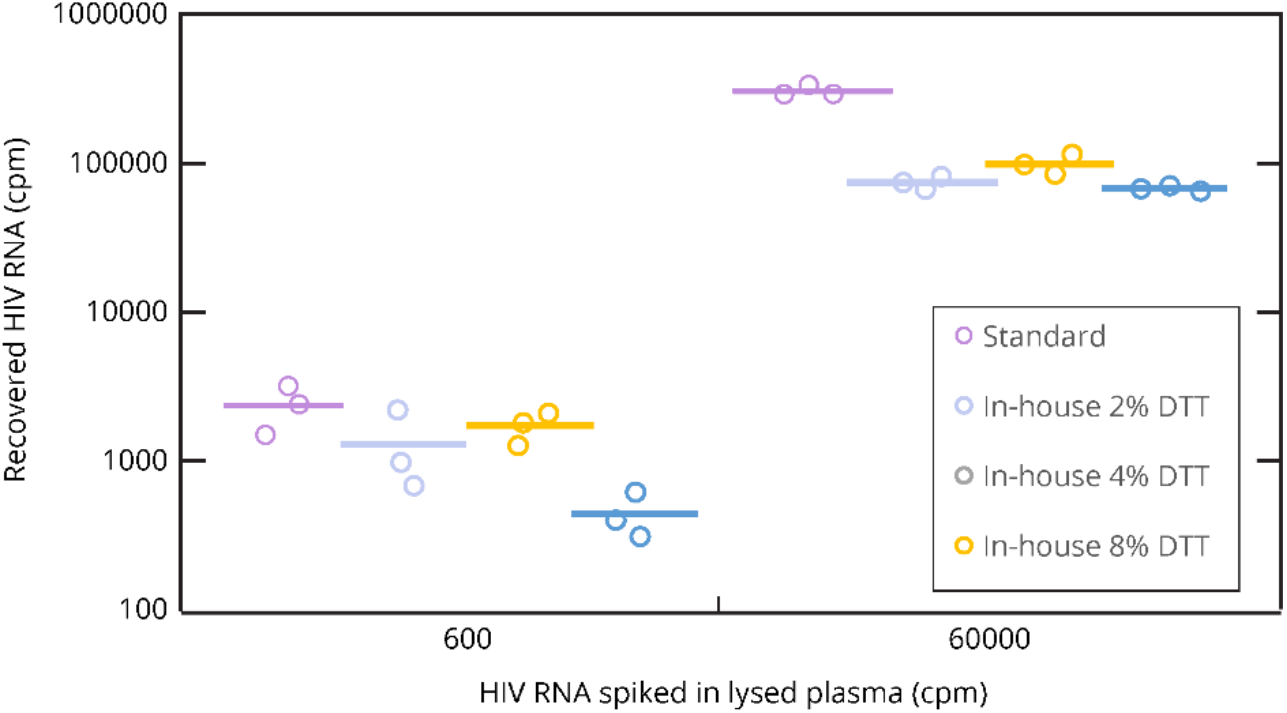
Effect of DTT concentrations in in-house lysis buffer (4M GuSCN, 10mM MES pH 5.5) The in-house RNA lysis buffer containing 2 - 8% (v/v) DTT (2M) or the standard lysis buffer (Qiagen ViralAmp kit) was used to lyse HIV negative human plasma. After 10min (Qiagen protocol) and 15min (in-house protocol) incubation, HIV RNA was spiked into the samples either at 0, 600, or 60000 copies/mL. The samples were then proceeded with the rest of the extraction protocol and analyzed using RT-qPCR. The Cq values from each condition were converted to recovered copies and plotted (mean±SE, *n = 3*).

**Supplementary figure 4.**
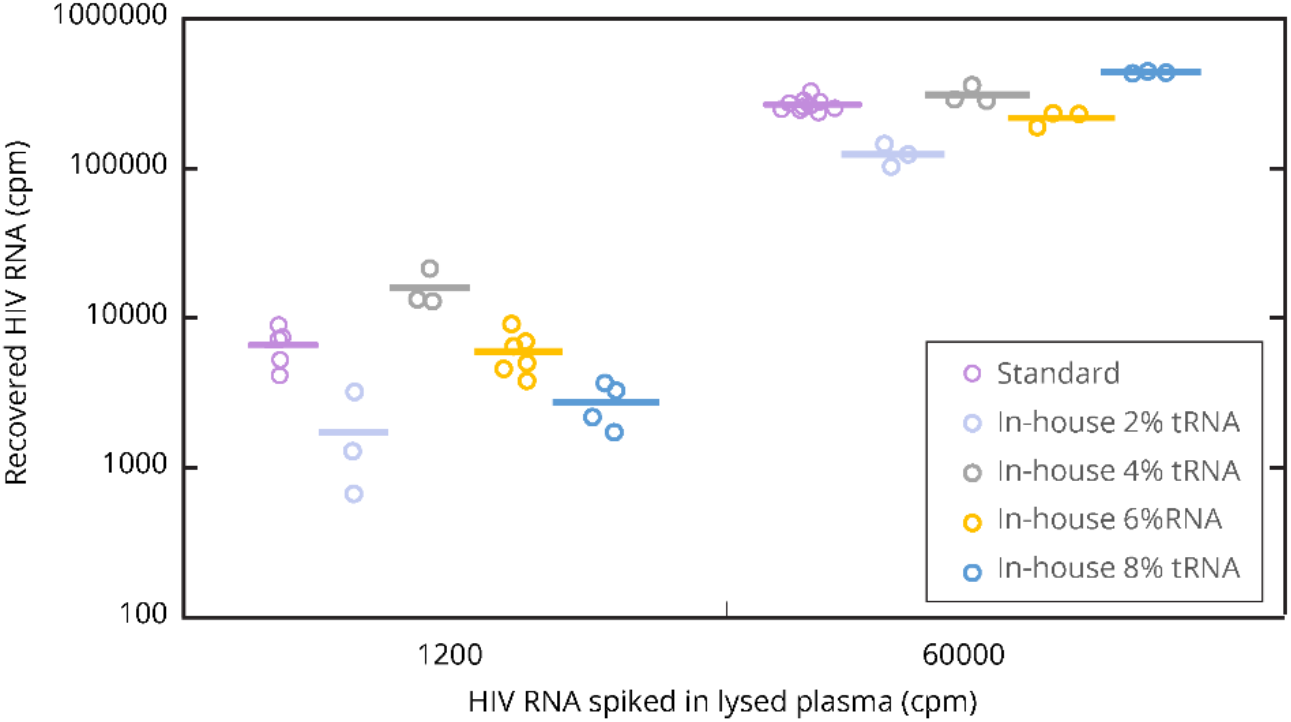
Effect of carrier RNA in in-house lysis buffer (4M GuSCN, 10mM MES pH 5.5, 4% DTT). The in-house RNA lysis buffer containing 2 - 8% (v/v) 1mg/mL tRNA or the standard lysis buffer (Qiagen ViralAmp kit) was used to lyse HIV negative human plasma. After 10min (Qiagen protocol) and 15min (in-house protocol) incubation, HIV RNA was spiked into the samples either at 0, 1200, or 60000 copies/mL. The samples proceeded with the rest of the extraction protocol and analyzed using RT-qPCR. The Cq values from each condition were converted to recovered copies and plotted (mean ± SE, *n = 3*).

Next, we integrate the new extraction method with the new RT-qPCR. Presence of carrier tRNA used in the in-house method can compete with the HIV RNA target during the RT-qPCR amplification, thus we conducted an experiment to determine optimal volume of the extracted RNA that should be added into each RT-qPCR reaction. Using 5µL extracted RNA (from 8%(v/v) tRNA lysis buffer) in 20 µLRT-PCR led to RT-PCR inhibition (green lines, **Supplementary figure 5a)**. Using 1%(v/v) of standard carrier RNA with the in-house buffer avoided inhibition (blue lines, **Supplementary figure 5a**) but differed from the standard Qiagen kit (red lines, **Supplementary figure 5a**) by about 2 cycles. When using 2µL extracted RNA in 20µL reaction, the inhibition from using 8µg/µL tRNA (green lines, **Supplementary figure 5b**) disappears, and recovery is better than when using 1ug/µL Qiagen carrier RNA (blue lines, **Supplementary figure 5b**), but is still not as high as the standard Qiagen kit (red lines, **Supplementary figure 5b**). The difference between in house method with 8%(v/v) tRNA and Qiagen kit is ∼1 cycle. Based on these results we have finalized our in-house protocol using 6%(v/v) tRNA in the in-house lysis buffer with 5µLextracted RNA in 20µLRT-PCR reaction.

**Supplementary figure 5.**
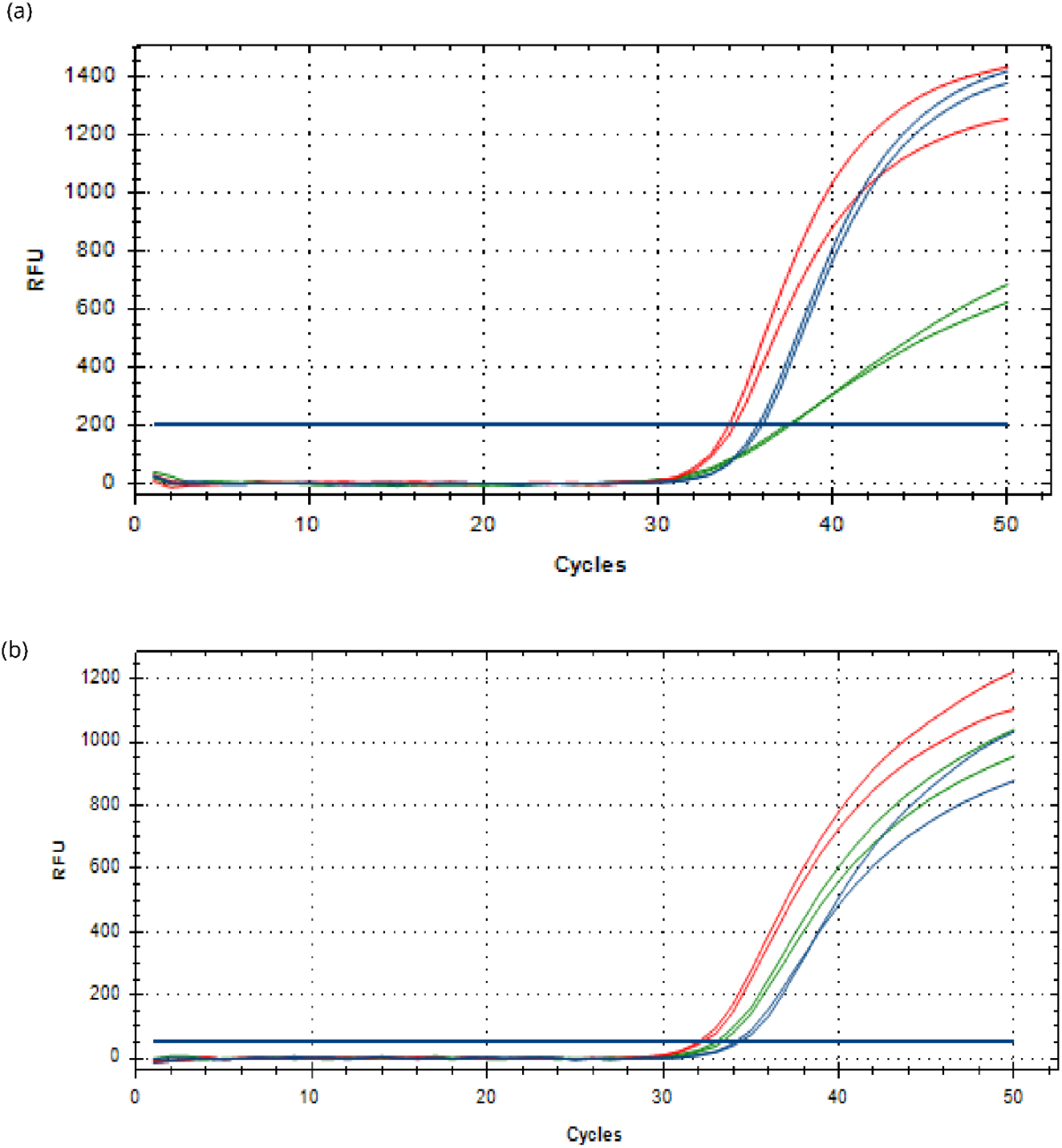
Impact of carrier tRNA concentration on RT-qPCR. (**a**) Amplification curves of 5µL extracted RNA (**b**) Amplification curves of 2µL extracted RNA from HIV RNA 1000copies/mL using either standard protocol (red lines), standard tRNA condition (1%) in in-house buffer (blue lines), or 8% tRNA in in-house buffer (green lines). Each condition was performed in duplicates and individually plotted.

**Supplementary figure 6.**
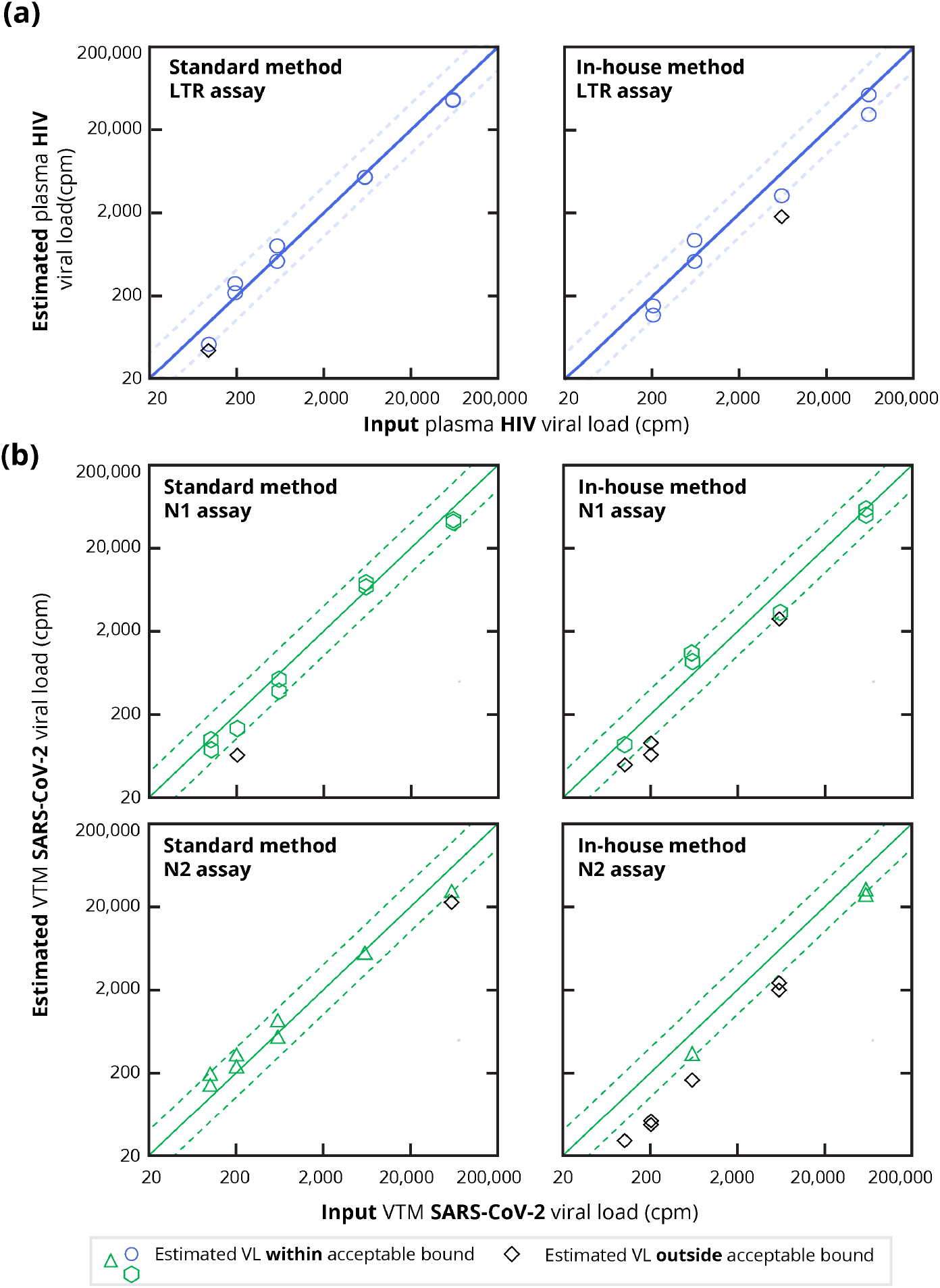
Correlation plots of measured VL from co-extraction of contrived plasma/NS. Measured VL of (**a**) HIV and (**b**) SARS-CoV-2 targets in contrived plasma samples (lysed plasma and spiked with synthetic HIV RNA and SARS-CoV-2 RNA at 0, 100, 200, 600, 6000, or 60000 copies/mL). All individual data are plotted. Diagonal lines represent 100% recovery with the dash lines indicating precise measurement bound of ±0.3 log10 (VL input, copies/mL).

**Supplementary figure 7.**
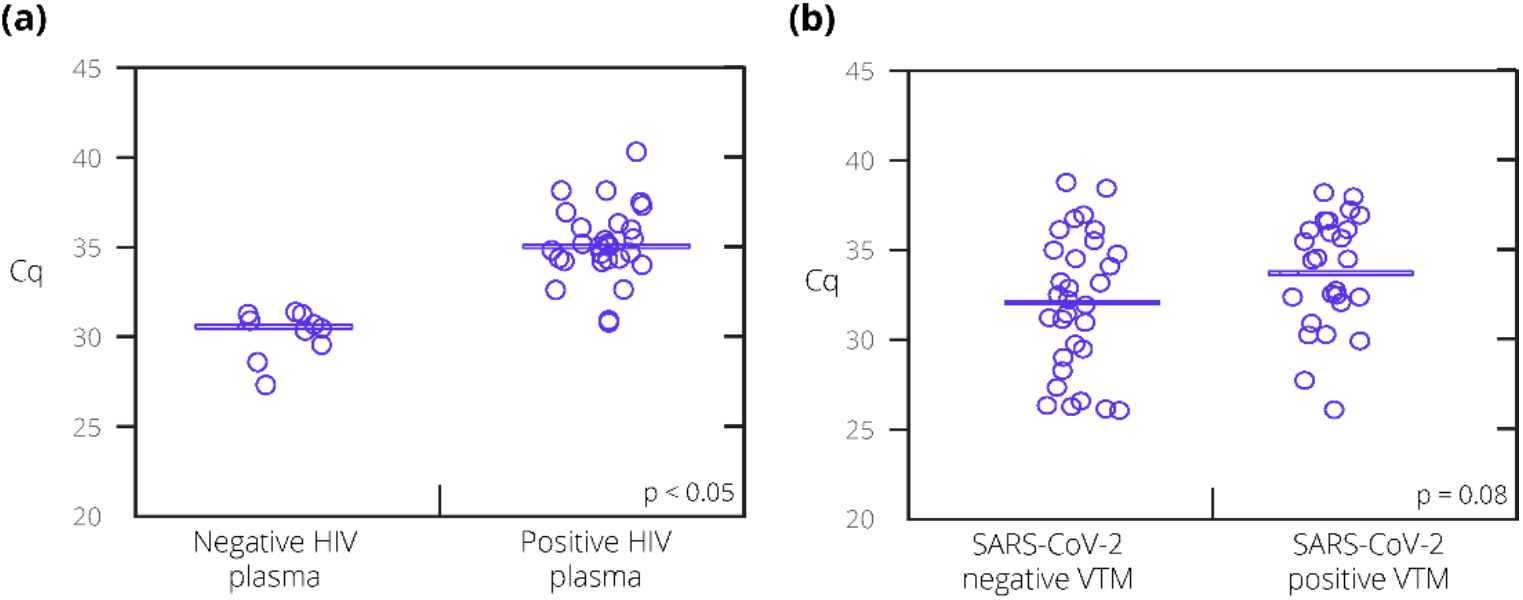
Levels of RP gene of clinical specimens. **(a)** Plasma. HIV-seronegative plasma were purchased from Innovative Research, Inc, MI, USA. HIV-seropositive plasma were collected from treatment naive individuals in Mexico City. **(b)** NS both SARS-CoV-2 positives and negatives were collected from individuals in the US with respiratory symptoms in the US. All specimens were stored at –80 °C until test. Averages of the positive and negative groups were assessed for statistical difference using Student’s t-test (two-tailed).

**Supplementary table 1.**
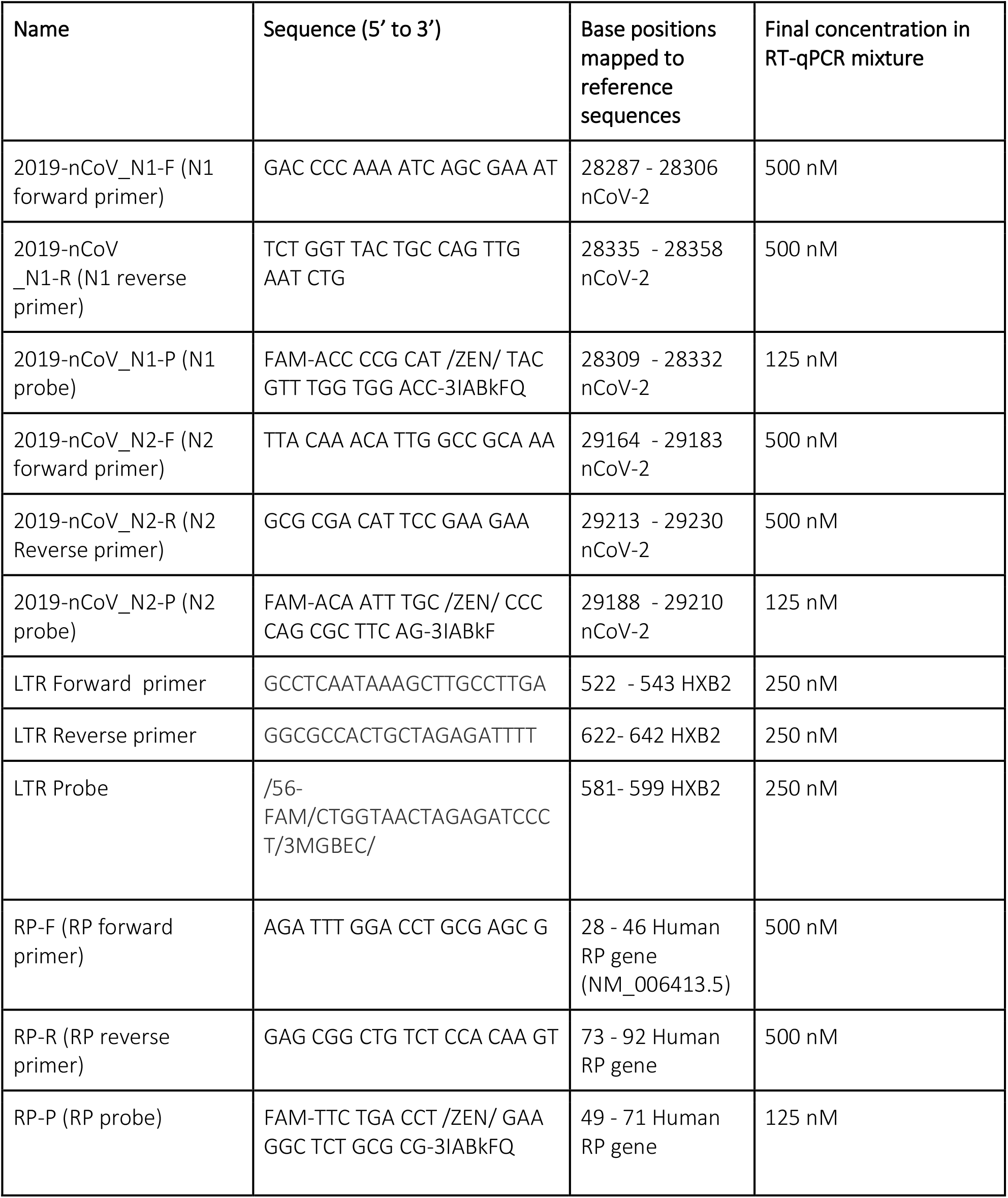
Primers and probe sequences nCoV-N control: severe acute respiratory syndrome coronavirus 2 isolate Wuhan-Hu-1, complete genome (GenBank: NC_045512.2)^[31, 42]^

**Supplementary table 2.** Cost estimate for the RNA extraction kit

| Consumables/reagents | Source | Unit | Cost |
| --- | --- | --- | --- |
| GuSCN | VWR, 97061-524 | 500g | \$252.96 |
| MES buffer | Sigma, 76039 | 1M, 0.5L | \$116 |
| Tris-HCl buffer | Fisher, 15568025 | 1M, 1L | \$95.25 |
| DTT | Promega, C4H10O2S2 | 5g | \$139.10 |
| Collection tubes | Qiagen, 19201 | 1000 tubes | \$166 |
| Spin column tubes | Epoch Life Science, 1920 | 250 tubes | \$100 |
| tRNA | Fisher, AM7119 | 5mg | \$95.25 |
| **Buffer (50mL stock)** |  |  |  |
| Lysis buffer | 4M GuSCN, 10mM MES | 50mL | \$13.66 |
| Wash buffer 1 | 1M GuSCN, 10mM Tris-HCl | 50mL | \$3.18 |
| Wash buffer 2 | 10mM Tris-HCl | 50mL | \$0.05 |
| **Cost/kit (140µL sample)** |  |  |  |
| Epoch column | | | \$0.40 |
| Collection tubes | | | \$0.51 (3 x \$0.17) |
| Lysis buffer | | | \$0.15 |
| tRNA | | | \$0.64 |
| DTT | | | \$0.10 |
| Wash buffer 1 | | | \$0.05 |
| Wash buffer 2 |  |  | negligible |
| **Total cost** | | | **\$1.85** |

## References

1. Adepoju P. Tuberculosis and HIV responses threatened by COVID-19. The lancet HIV 2020; 7(5):e319–e320.

2. Shiau S, Krause KD, Valera P, Swaminathan S, Halkitis PN. The Burden of COVID-19 in People Living with HIV: A Syndemic Perspective. AIDS and behavior 2020; 24(8):2244–2249.

3. Darcis G, Vaira D, Moutschen M. Impact of coronavirus pandemic and containment measures on HIV diagnosis. Epidemiology and infection 2020; 148:e185.

4. Kindzeka ME. Cameroon’s HIV/AIDS Patients Shirk Hospitals for Fear of COVID-19. In; 2020.

5. Bhaskaran K, Rentsch CT, MacKenna B, Schultze A, Mehrkar A, Bates CJ, et al. HIV infection and COVID-19 death: a population-based cohort analysis of UK primary care data and linked national death registrations within the OpenSAFELY platform. The Lancet HIV 2021; 8(1):e24–e32.

6. Hadi YB, Naqvi SFZ, Kupec JT, Sarwari AR. Characteristics and outcomes of COVID-19 in patients with HIV: a multicentre research network study. AIDS 2020; 34(13):F3–F8.

7. Osibogun A, Balogun M, Abayomi A, Idris J, Kuyinu Y, Odukoya O, et al. Outcomes of COVID-19 patients with comorbidities in southwest Nigeria. PloS one 2021; 16(3):e0248281.

8. Mellor MM, Bast AC, Jones NR, Roberts NW, Ordóñez-Mena JM, Reith AJM, et al. Risk of adverse coronavirus disease 2019 outcomes for people living with HIV. Aids 2021; 35(4):F1–f10.

9. Mirzaei H, McFarland W, Karamouzian M, Sharifi H. COVID-19 Among People Living with HIV: A Systematic Review. AIDS and behavior 2021; 25(1):85–92.

10. Tesoriero JM, Swain C-AE, Pierce JL, Zamboni L, Wu M, Holtgrave DR, et al. COVID-19 Outcomes Among Persons Living With or Without Diagnosed HIV Infection in New York State. JAMA Network Open 2021; 4(2):e2037069–e2037069.

11. Ambrosioni J, Blanco JL, Reyes-Urueña JM, Davies M-A, Sued O, Marcos MA, et al. Overview of SARS-CoV-2 infection in adults living with HIV. The Lancet HIV 2021; 8(5):e294–e305.

12. Susman E. Acute HIV Cases Turn Up in COVID Screening — Similar symptoms result in surge in finding new infections at one institution. In; 2020.

13. Karim F, Moosa M, Gosnell B, Cele S, Giandhari J, Pillay S, et al. Persistent SARS-CoV-2 infection and intra-host evolution in association with advanced HIV infection. medRxiv 2021:2021.2006.2003.21258228.

14. HIV.gov. Coronavirus (COVID-19) and People with HIV. In; 2021.

15. World Health Organization. WHO: access to HIV medicines severely impacted by COVID-19 as AIDS response stalls. In; 2020.

16. Chenneville T, Gabbidon K, Hanson P, Holyfield C. The Impact of COVID-19 on HIV Treatment and Research: A Call to Action. International journal of environmental research and public health 2020; 17(12).

17. Prebensen C, Myhre PL, Jonassen C, Rangberg A, Blomfeldt A, Svensson M, et al. Severe Acute Respiratory Syndrome Coronavirus 2 RNA in Plasma Is Associated With Intensive Care Unit Admission and Mortality in Patients Hospitalized With Coronavirus Disease 2019. Clinical Infectious Diseases 2020; 73(3):e799–e802.

18. Colagrossi L, Antonello M, Renica S, Merli M, Matarazzo E, Travi G, et al. SARS-CoV-2 RNA in plasma samples of COVID-19 affected individuals: a cross-sectional proof-of-concept study. BMC Infectious Diseases 2021; 21(1):184.

19. Veyer D, Kernéis S, Poulet G, Wack M, Robillard N, Taly V, et al. Highly Sensitive Quantification of Plasma Severe Acute Respiratory Syndrome Coronavirus 2 RNA Sheds Light on its Potential Clinical Value. Clinical Infectious Diseases 2020.

20. Prebensen C, Myhre PL, Jonassen C, Rangberg A, Blomfeldt A, Svensson M, et al. Severe Acute Respiratory Syndrome Coronavirus 2 RNA in Plasma Is Associated With Intensive Care Unit Admission and Mortality in Patients Hospitalized With Coronavirus Disease 2019. Clinical Infectious Diseases 2020.

21. New England Biolabs I. Protocol for Standard RNA Synthesis. In; 2021.

22. New England Biolabs I. Monarch® RNA Cleanup Kit (50 μg). In; 2021.

23. Panpradist N, Wang Q, Ruth PS, Kotnik JH, Oreskovic AK, Miller A, et al. Simpler and faster Covid-19 testing: Strategies to streamline SARS-CoV-2 molecular assays. EBioMedicine 2021; 64.

24. Panpradist N, Wang Q, Ruth PS, Kotnik JH, Oreskovic AK, Miller A, et al. Simpler and faster Covid-19 testing: Strategies to streamline SARS-CoV-2 molecular assays. EBioMedicine 2021; 64:103236.

25. Panpradist N, Beck IA, Ruth PS, Ávila-Ríos S, García-Morales C, Soto-Nava M, et al. Near point-of-care, point-mutation test to detect drug resistance in HIV-1: a validation study in a Mexican cohort. AIDS 2020; 34(9).

26. QIAGEN. QIAamp® Viral RNA Mini Handbook. In. HB-0354-0007 ed; 2020.

27. Drain PK, Dorward J, Bender A, Lillis L, Marinucci F, Sacks J, et al. Point-of-Care HIV Viral Load Testing: an Essential Tool for a Sustainable Global HIV/AIDS Response. Clinical microbiology reviews 2019; 32(3).

28. U.S. National Institute of Health. GLOSSARY of HIV/AIDS-Related Terms. In. 9th ed; 2021.

29. World Health O. Consolidated guidelines on the use of antiretroviral drugs for treating and preventing HIV infection: recommendations for a public health approach. 2nd ed ed. Geneva: World Health Organization; 2016.

30. U.S. Centers for Disease Control and Prevention. CDC 2019-Novel Coronavirus (2019-nCoV) Real-Time RT-PCR Diagnostic Panel. In; 2020.

31. Arvold ND, Ngo-Giang-Huong N, McIntosh K, Suraseranivong V, Warachit B, Piyaworawong S, et al. Maternal HIV-1 DNA load and mother-to-child transmission. AIDS patient care and STDs 2007; 21(9):638–643.

32. U.S. Centers for Disease Control and Prevention. CDC 2019-Novel Coronavirus (2019-nCoV) Real-Time RT-PCR Diagnostic Panel (Instructions for Use). In. 6 ed: U.S. Food & Drug Administration; 2020.

33. Boom R, Sol CJ, Salimans MM, Jansen CL, Wertheim-van Dillen PM, van der Noordaa J. Rapid and simple method for purification of nucleic acids. Journal of Clinical Microbiology 1990; 28(3):495–503.

34. Padhye VV, York C, Burkiewicz A. Nucleic acid purification using silica gel and glass particles. In; 1993.

35. Bernhardt HS, Tate WP. Primordial soup or vinaigrette: did the RNA world evolve at acidic pH? Biology direct 2012; 7:4.

36. Escobar MD, Hunt JL. A cost-effective RNA extraction technique from animal cells and tissue using silica columns. Journal of biological methods 2017; 4(2):e72.

37. Mommaerts K, Sanchez I, Betsou F, Mathieson W. Replacing β-mercaptoethanol in RNA extractions. Analytical Biochemistry 2015; 479:51–53.

38. Oreskovic A, Brault ND, Panpradist N, Lai JJ, Lutz BR. Analytical Comparison of Methods for Extraction of Short Cell-Free DNA from Urine. The Journal of Molecular Diagnostics 2019; 21(6):1067–1078.

39. Panpradist N, Beck IA, Vrana J, Higa N, McIntyre D, Ruth PS, et al. OLA-Simple: a software-guided HIV-1 drug resistance test for low-resource laboratories. EBiomedicine 2019; In press.

40. Organization WH. WHO in-house assay. In; 2020.

41. Pau CP, Wells SK, Rudolph DL, Owen SM, Granade TC. A rapid real-time PCR assay for the detection of HIV-1 proviral DNA using double-stranded primer. J Virol Methods 2010; 164(1-2):55–62.

42. U.S. Centers for Disease Control and Prevention. CDC 2019-Novel Coronavirus (2019-nCoV) Real-Time RT-PCR Diagnostic Panel For Emergency Use Only. In: U.S. Food and Drug Administration; 2020.

43. Byrnes SA, Gallagher R, Steadman A, Bennett C, Rivera R, Ortega C, et al. Multiplexed and Extraction-Free Amplification for Simplified SARS-CoV-2 RT-PCR Tests. Analytical chemistry 2021; 93(9):4160–4165.

44. Oreskovic A, Panpradist N, Marangu D, Ngwane MW, Magcaba ZP, Ngcobo S, et al. Diagnosing pulmonary tuberculosis using sequence-specific purification of urine cell-free DNA. J Clin Microbiol 2021.

45. Zhao Z, Cui H, Song W, Ru X, Zhou W, Yu X. A simple magnetic nanoparticles-based viral RNA extraction method for efficient detection of SARS-CoV-2. bioRxiv 2020:2020.2002.2022.961268.

46. Iwase SC, Miyazato P, Katsuya H, Islam S, Yang BTJ, Ito J, et al. HIV-1 DNA-capture-seq is a useful tool for the comprehensive characterization of HIV-1 provirus. Scientific Reports 2019; 9(1):12326.

